# Genome-wide analysis of individual coding variants and HLA-II-associated self-immunopeptidomes in ulcerative colitis

**DOI:** 10.1101/2023.03.22.23286498

**Authors:** Mareike Wendorff, Hesham ElAbd, Frauke Degenhardt, Marc Höppner, Florian Uellendahl-Werth, Eike M. Wacker, Lars Wienbrandt, Simonas Juzenas, Regeneron Genetic Center, Tomas Koudelka, David Ellinghaus, Petra Bacher, Andreas Tholey, Matthias Laudes, Malte Ziemann, Bernd Bokemeyer, Stefan Schreiber, Tobias L. Lenz, Andre Franke

## Abstract

Genome wide association studies contributed to a better understanding of the etiology of inflammatory bowel disease (IBD). While over 240 genetic associations with IBD have since been identified, functional follow-up studies are still in their infancy with the overall pathogenesis of IBD remaining unsolved. E.g., a functional understanding of the genetic association between the human leukocyte antigen (HLA) region and ulcerative colitis (UC) – one subtypes of IBD – is still lacking. Here, we analyzed whether an autoimmune reaction involving the HLA class II proteins HLA-DQ and -DR, both being strongly associated with UC, could be a disease trigger or driver. To this end, genotype data derived from whole exome sequencing and genome-wide SNP array data of 863 German UC patients as well as 4,185 healthy controls were analyzed. Association analyses identified novel variants in the *NOD2* and *SNX20* genes to be linked with UC and confirmed known HLA allele associations. Employing the genetic data, we generated patient-specific self-immunopeptidomes and *in sili*co predicted HLA-peptide binding. Peptidome-wide association analyses of peptide binding preferences in a set of candidate proteins yielded significant associations with 234 specific peptides. Interestingly, none of those peptides showed a differential presence in case and control samples. The disease-associated candidate peptides predicted to be presented by risk HLA proteins contained predominantly aromatic amino acids. In contrast, protective HLA proteins were predicted to bind peptides enriched in acidic amino acids. In summary, we present a proof-of-concept immunogenetic analysis that contributes to a better understanding of the HLA in UC.

## Introduction

Although about 0.3% of people in the industrialized countries suffer from inflammatory bowel disease (IBD)^1^, the etiology of the diseases is still unclear. Different studies have correlated the disease with environmental^2, 3^ and genetic factors^4–7^. For some of the known factors, a concrete role in the pathogenesis of IBD has been identified, in other cases the impact on the disease remains unknown. Interestingly, the major histocompatibility complex region (MHC), which shows the strongest genetic association with the disease, still has an enigmatic role in IBD^8^. The genetic association at this locus differs between the two main subtypes of IBD, Crohn’s disease (CD) and ulcerative colitis (UC)^8, 9^. Especially in UC, the MHC class II genes are highly associated with the disease in Caucasians (Goyette *et al.*^9^) and across different ancestries (Degenhardt *et al.*^10^). Though detailed analysis showed a consistent genetic profile, the biology behind the associations remains unsolved. MHC class II proteins mainly present peptides derived from extracellular proteins to CD4-positive T cells, which may then elicit an immune response in the host. The classical MHC class II genes in humans are the human leukocyte antigen (HLA) genes HLA-*DPA1*, -*DPB1*, -*DQA1*, -*DQB1*, -*DRA*, -*DRB1* and depending on the (*DRB1*-related) haplotype possibly one of HLA-*DRB3*, -*DRB4* and -*DRB5*. Except for HLA-*DRA*, which is nearly invariable regarding its protein sequence, all other classical HLA proteins are highly polymorphic. Several different processes of how the HLA may contribute to the inflammation in IBD have been discussed in the literature, most of those are based on the structural differences in the peptide binding pocket and the related differences in antigen recognition^8^. Differences among HLA alleles and their binding preferences have already been discussed in Goyette *et al.*^9^ and Degenhardt *et al.*^10^. Here, we dig deeper into the *autoimmune hypothesis* which refers to an immune reaction triggered by HLA-presentation of a host’s self-peptide^8^.

For this purpose, we analyzed genotype data derived from next generation sequencing (NGS)-based whole exome-sequencing (WES) and genotyping based on Illumina’s Global Screening Array (GSA) as well as imputed HLA allele information of 5,048 German individuals for genetic association with UC (**Figure 1, Supplementary Table 1**). This data is unprecedented in its resolution of individual-level coding variation.

**Figure 1:**
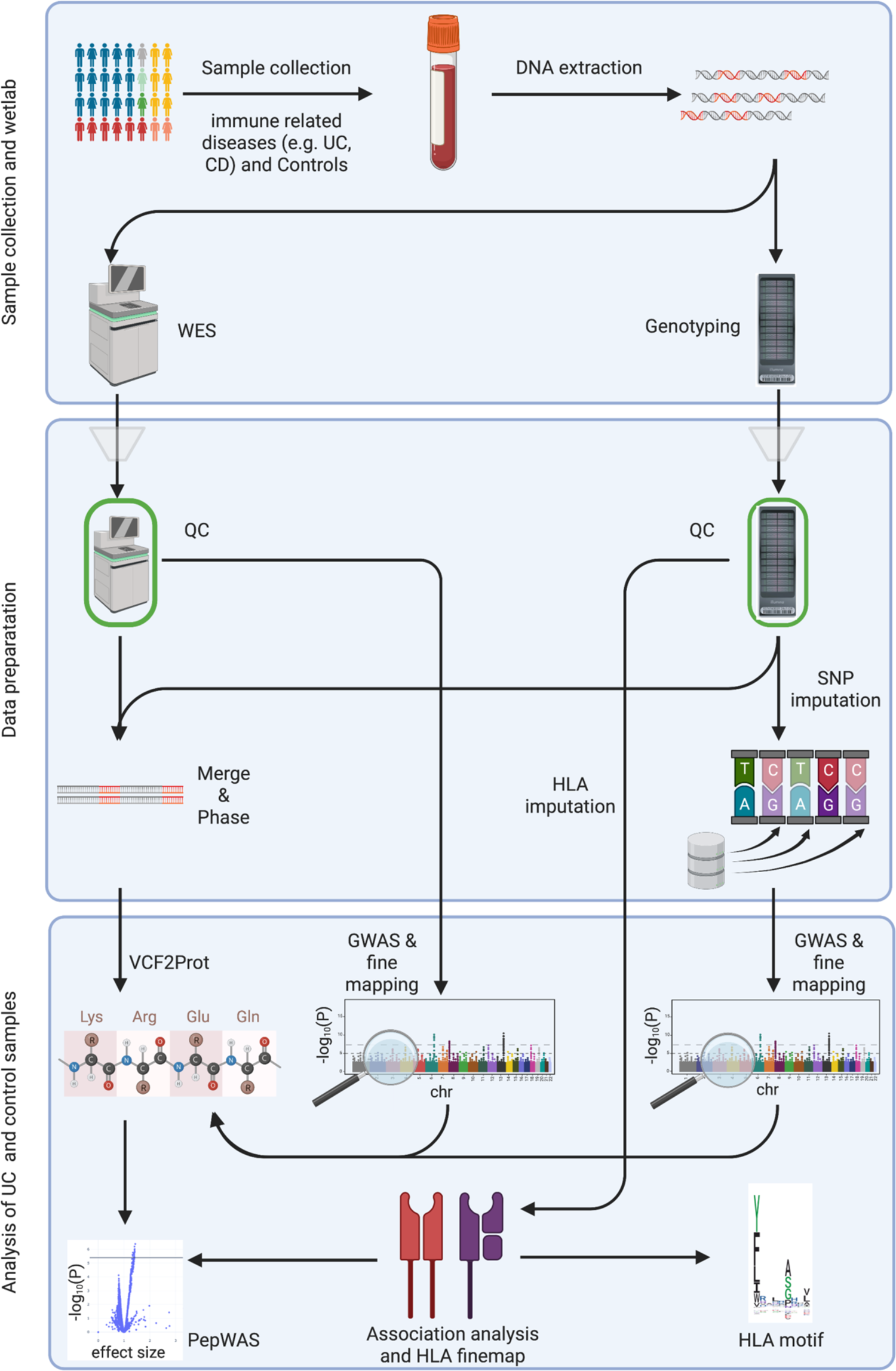
Graphical abstract of the current study. The first box summarizes the sample collection and wetlab part. The second box shows the data preparation. The third box lists the different performed analyses. UC: ulcerative colitis, CD: Crohn’s disease, DNA: Deoxyribonucleic acid, QC: quality control, SNP: single-nucleotide polymorphism, HLA: human leucocyte antigen, GWAS: genome-wide association study, PepWAS: peptidome-wide association study. This Figure was created with BioRender.com.

Based on this exceptionally high-resolution genotype data, we first performed a genome-wide association analysis (GWAS) as well as a fine-mapping of the HLA region. Subsequently, we took advantage of the individual WES data and performed a peptidome-wide association analysis (PepWAS)^11^ based on personalized proteomes, focusing on self-peptides from a disease-relevant set of human proteins (**Supplementary Table 2**), to identify candidate self-peptides with a differential likelihood of presentation in patients and controls. The personalized proteomes were generated by translating the per-patient observed nucleotide variations from the WES data into a collection of personalized protein sequences. Further, we checked the peptides identified through the PepWAS for mutations in the coding DNA sequence leading to the according peptide or another amino acid sequence.

## Results

### Comparison of exome data and imputed genotyping data

The exome data is expected to include novel or rarely described coding variants. In a first step we therefore compared how many exome variants are not included in our imputed SNP (single-nucleotide polymorphism) genotyping array dataset. The comparison revealed that half of the variants discovered by exome sequencing are not detected in the imputed data. Most of those variants are very rare (minor allele frequency (MAF)<0.1%). Of the variants with a higher allele frequency about 11% are unique to the exome data. 6.3% of the common exome variants (MAF>5%) are still not imputed.

The fraction of variants specific to the exome data varies not only with the allele frequency but also with the type of mutation. E.g., for missense variants the most common variants were imputed and only 2.8% of variants were specific for the exome dataset but the fraction of overlap is lower for InDels.

### GWAS of imputed genotyping data

For the summary statistics of the imputed genotyping data, a genomic inflation factor lambda of 0.995 was calculated based on the data excluding the HLA region (**Supplementary Figure 1**). This means no population stratification is expected to cause false positive hits. The Manhattan plot of the analysis is shown in **Supplementary Figure 2**. Overall, 1,713 of the imputed genotyping markers with a minor allele frequency of at least 1% passed the suggestive significance threshold P-value<10^-5^ and 975 of those reached genome-wide significance. The at least suggestively significant markers were assigned to 26 loci, considering two variants as belonging to one locus if their physical distance in GRCh38 is below 150 kbp, overlapping largely with results from LD-based clumping (P-value<0.00001) but allowing for a combination of larger association signals such as the HLA class II with 24 clumped signals calculated by PLINK. Of these loci 19 main variants were supported by variants in LD (**Supplementary Table 3**; **Supplementary Figures 3-21**). Three of those loci contained genome wide significant variants: 1p36.13 (*OTUD3,* **Supplementary Figure 5**), 6p21.32 (*HLAII,* **Supplementary Figure 11**), and 5p14.3 (no specific gene, **Supplementary Figure 9**). In agreement with previously published GWAS^7^, our dataset shows the main significant genetic association in the HLA region on chromosome 6 with the main peak being in the region of the classical HLA class II genes. In addition, *OTUD3* is one of the strongest associated loci reported previously in the literature for UC^7^. 5p14.3 (**Supplementary Figure 9**) was not reported previously and does not replicate in the publicly available IIBDGC dataset (available through RICOPILI)^30^. The main signal rs2937516 (P-value=1.62×10^-8^, OR=0.68 [0.60–0.78]) is located on chr5:18748431 between the genes *LINC02100* (chr5:18,514,857-18,746,202) and the pseudogene *UBE2V1P12* (chr5:18,886,622-18,887,095).

Next to the three genome-wide significant loci, we identified 16 suggestively significant loci (**Supplementary Table 3**): 5p13.1 (*PTGER4*, **Supplementary Figure 10**)^7^*, 10q24.2 (NKX2-3*, **Supplementary Figure 14**)^7^ and 22q13.1 (*PDGFB - RPL3*, **Supplementary Figure 21**)^7^ as well as 6p21.33 (*HLAI,* **Supplementary Figure 11**) all of which being previously reported to be associated with UC (NHGRI-EBI GWAS catalog^29^). An intriguing novel association signal for UC was detected at 16q12.1 (*NOD2 and SNX20*, **Supplementary Figure 17**), a locus known previously to be associated only with Crohn’s disease^7,54^. A detailed follow up-analysis of these variants in the context of UC is described in the paragraph *SNX20/NOD2 association in ulcerative colitis and Crohn’s disease.* 12q24.13-q24.21 (*RBM19*, rs3782449, P-value= 8.90×10^-6^, OR=0.73 [0.64-0.84], **Supplementary Figure 16**) and 4p12 (rs113429955, P-value=5.51×10^-6^, OR=1.81 [1.40-2.33], **Supplementary Figure 8**) have not been previously described but were replicated using the IIBDGC dataset with replication-P-values of 0.00738 and 0.045, respectively. For 12q24.13-q24.21 (*RBM19*), an association with ocular manifestation in IBD was described by others before^55^. The LD between the T allele rs4766697, the variant associated with ocular manifestation and the A allele rs3782449, our protective lead variant, is with R^2^ of 0.0058 very low but the D’ is 1. Therefore, all T of rs4766697 (allele frequency of 2.2 %, P-value=0.64, OR=0.92 [0.64-1.32] in our data) are most likely located on the haplotype of the reference allele A of rs3782449 (allele frequency 79.3 %). The two-sided Fisher’s Exact test on the contingency table of the dosages yielded a P-value of 1.27×10^-7^.

The region of the signal at 9q22.2 (*UNQ6494*, rs36147380, P-value=7.4×10^-6^, OR=0.74 [0.65-0.84], **Supplementary Figure 13**) was not covered in the GRCh37 build and therefore no lookup in the IIBDGC GWAS data was possible. The remaining eight loci did neither replicate in the IIBDGC GWAS dataset nor were they listed in the NHGRI-EBI GWAS catalog, but four of these loci had additional links to UC. Firstly, *TNFRSF8*, also called *CD30L*, at 1p36.22 (rs72641067, P-value=7.4×10^-3^, OR=1.47 [1.25-1.77], **Supplementary Figure 3**) is an (auto-)immune relevant gene and the gene coding for the corresponding ligand gene *TNFSF8* has been described as associated with CD^7,56^. Secondly, the gene *TPRG1* (3q28, chr3:188,947,214-189,325,304) was found to be associated in the IBD GWAS from de Lange *et al.*^5^, but the location of the signal was slightly different. Our lead SNP rs73184427 is located at chr3:189,167,658 (P-value=7.11×10^−06^; OR=1.56 [1.28–1.89], **Supplementary Figure 7**), while de Lange *et al.* described rs56116661 at chr3:188,683,372, annotated to the *LPP* gene, to be associated with CD (P-value=5.67×10^−10^; OR=1.14 [1.10–1.18])^5^. The R^2^ and the D’ between the variants are with respectively 0.00092 and 0.18 low and the association P-value of the previously described variant rs56116661 in UC patients vs. controls is 0.0067 (OR = 0.82 [0.72–0.95]). Thirdly, the gene *CTCF* (16q22.1, rs117327757, chr16:67627037, P-value = 8.6×10^-6^, OR = 1.8 [1.39-2.33], **Supplementary Figure 18**) is known to influence the expression of TNF^57^. Fourthly, *LYPD5* (19q13.31, rs364691, chr19:43804850, P-value = 1.6e10-6, OR = 1.35 [1.2-1.53]) was reported by Taman *et al.* as upregulated in treatment-naïve UC patients^58^.

### Associations in the whole exome data

The genomic inflation factor lambda for the exome data was determined to be 1.005 (**Supplementary Figure 22**). Overall, five loci showed at least suggestive associations (**Supplementary Figure 23-28, Supplementary Table 3**). In the whole exome data only the HLA-II region was found as a genome-wide significant signal with LD support (**Supplementary Figure 25**). The intronic *PGAM5* variant rs7973452 at 12p13.2 is LD-supported and reaches suggestive significance (P-value=8.47×10^-6^; OR=1.34 [1.18-1.53], **Supplementary Figure 27**). PGAM5 is known to regulate antiviral responses^59^. Three additional loci reached suggestive significance, none of these were supported by variants in LD, even when including low frequency variants. One of those is the 1p36.13 locus with *OTUD3,* also identified in the imputed genotyping dataset, with the intronic variant rs773646156 (chr1:19890367; P-value=4.53×10^−7^; OR=1.33 [1.19–1.48], **Supplementary Figure 24**). Second, the bitter taste receptor gene *TAS2R43*, with the missense variant rs200922417 (chr12:11092088; P-value=6.17×10^−6^; OR=1.92 [1.45–2.54]; 48L>48V, **Supplementary Figure 26**), is a suggested target for UC treatment as it influences *CLCA1*^60^. This variant is not well covered in the imputed genotyping data and the gene was not distinguished from *TAS2R45* in genome build GRCh37. The fifth association in the exome data, the third without LD support, is the synonymous variant rs755163625 on 22q11.21 in the gene *SCARF2* (chr22:20429374; P-value=9.2×10^-6^; OR=2.37 [1.62-3.47], **Supplementary Figure 28**). Coding mutations within *SCARF2* were previously described as responsible for the Van Den Ende-Gupta Syndrome, an extremely rare autosomal-recessive disorder characterized by distinctive craniofacial features^61^.

### SNX20/NOD2 association in ulcerative colitis and Crohn’s disease

*NOD2*, also formerly known as *IBD1*^4^ in linkage studies, was previously described to be a CD-specific disease gene. Here, we identified an association with UC. To investigate this further, we performed additional lookups and calculated the genetic association for this region also for the available CD data (4,097 CD cases and 4,185 controls as in UC analysis).

Our main UC association in the genotyping dataset rs139397276 (P-value=1.3×10^−6^; OR=3.74 [2.19–6.37]; MAF_controls_=0.9%; MAF_cases_=2.4%) was located at chr16:50666737 (**Supplementary Figure 17 and 29**). In addition, an association was identified with rs61736932 in the exome dataset (OR=4.22 [2.36, 7.54], P-value=1.15×10^-6^, MAF=1%, note: MAF is with 0.00990 slightly below 1% therefore it was not described above) at chr16:50711744. Both rs139397276 and rs61736932 are located closer to the centromere as compared to the established CD susceptibility variants rs2066844, rs2066845, and rs2066847 (all of them have an R^2^=0 with the lead variant rs139397276). The variant rs139397276 is in the *SNX20* gene and the calculated credible set includes four additional variants with a probability above 1%, including the *NOD2* exome variant rs61736932 (OR=4.47 [2.43, 8.25], P-value=1.60×10^-6^, MAF=1%). Of the remaining three, two are in introns of the *NOD2* genes and one in an intron of *SNX20.* All five variants reach nominal significance in the association analysis (**Supplementary Table 3**, **Supplementary Figure 29**) and have a frequency around 1%. None of the variants changes the amino acid sequence of the NOD2 protein. While the previously described CD variants have an impact on the leucine rich repeat region (LRR) of the NOD2 protein, our variants are in the nucleotide binding region and in the 3’-UTR region of *SNX20* (**Supplementary Figure 29**). The IIBDGC dataset supported our finding of rs139397276 with a P-value of 2.6×10^-4^ and an OR of 1.36 [1.15-1.60]. We further queried the data of Lesage, 2002^62^ and of the IBD Exomes Browser^63^ for validation purposes. The data on the IBD Exomes Browser reports an overall P-value of 0.023 and an OR of 1.31 [1.07, 1.60] for rs61736932 in UC versus healthy controls. In the Non-Finnish European batch, the P-value is 6.78×10^-4^ with an OR of 1.93. Lesage reported with 1.9% (6 of 318) in UC patients a lower MAF than in their controls with 2.4% (5 of 206). Therefore, the data of Lesage and colleagues does not support the association we identified.

### Gene-based analysis

The gene-based analysis using SAIGE-GENE^35^ on the exome data with an AF<1% resulted in no significant gene if correcting for the number of analyzed genes (P-value < 2.61×10^-6^). We here describe the three genes with the lowest P-values. Those genes are *NOD2*, *KCNK3* and *PSORS1C1*.

*NOD2* was also identified in the single variant association of the imputed data. The same variant rs61736932 is responsible for the association signal of the gene-based test (P-value_SKAT-O_ = 3.47×10^-5^, P-value_SKAT_ = 1.21×10^-5^, P-value_Burden_ = 0.915). The variant was not identified in the single variant analysis as the allele frequency in the exome data was slightly below 1%. The Burden test result, with very low significance, suggests that the other rare variants have different directions of effect. Of the 13 included variants with an allele frequency between 0.1% and 1% only three other variants were more common in patients than in controls.

The gene *KCNK3* was previously not associated with IBD, of the three performed gene-based tests SKAT has the lowest P-value (P-value_SKAT-O_ = 1.10×10^-4^, P-value_SKAT_ = 5.17×10^-5^, P-value_Burden_ = 3.96×10^-2^). The signal is mainly based on the intronic variant rs926416351 with an allele frequency of 0.0059 (P-value = 4.07 ×10^-6^). In the very similar gene *KCNK9* a copy number variation (CNV) associated with UC was identified by Saadati *et al.*^64^. A recent study tested the effect of *KCNK9* and *KCNK3* knockout in a mouse model^65^. They figured out that the absence of one gene increases the expression of the other gene. A *KCNK3* knockout lead to a beneficial outcome in DSS-induced colitis with less mitochondrial damage and apoptosis, while the increased expression of *KCNK3* did not prevent apoptosis after DSS exposure.

The third gene *PSORS1C1* (P-value_SKAT-O_ = 3.04×10^-5^, P-value_SKAT_ = 4.27×10^-5^, P-value_Burden_ = 9.49×10^-5^) is located close to the HLA class I region. The two most common variants included in the gene-based analysis (rs118016068 and rs117114042) were also significantly associated in the publicly available RICOPILI^30^ dataset (see also **Supplementary Table 3**).

### Power analysis of genome wide associations in ulcerative colitis

Other well-known loci associated with UC like *IL23R*, *LINC02132* and *IL10* failed to show nominal significance but show the trend as reported in previous GWAS studies. The expected statistical power to reproduce those loci with a P-value of <10^-5^ is below 0.75 (**Supplementary Figure 30**). All previously reported loci with a greater suggested power were only reported in studies with samples of Asian populations and therefore could be population-specific findings.

### HLA association in UC

We imputed HLA alleles and amino acids for HLA-*A,-B*, -*C*, -*DPA1*, -*DPB1*, -*DQA1*, -*DQB1*, - *DRB1*, -*DRB3*, -*DRB4* and -*DRB5* at full 2-field level. Overall, we observed 234 different 2-field alleles in our UC and control data set. For the single loci, between 6 (HLA-*DPA1*) and 62 (HLA-*B*) different alleles were imputed into our samples. In consistency with previous studies of the HLA in UC^9,10^ the main association signal was in the locus containing HLA-*DR* and HLA-*DQ* (**Supplementary Figure 31**). Overall, four 2-field alleles were associated at genome-wide significance (*DQB1*\*06:02 (P-value=2.68×10^-10^, OR=1.58 [1.37-1.82]), *DRB1*\*15:01 (OR=1.56 [1.36-1.80], P-value=6.61×10^-10^), *DRB5*\*01:01 (OR=1.56 [1.35-1.80], P-value=8.40×10^-10^), *DRB4*\*01:03 (OR=0.66 [0.57-0.76], P-value=6.80×10^-9^)), and 3 additional 2-field alleles were nominally significant (5×10^-8^ < P-value < 1×10^-5^: *C*\*12:02, *B*\*52:01, *DQA1*\*01:02) (**Supplementary Table 4**). The strongest association from a SNP data was observed for the intronic SNP rs6927022 located in HLA-*DQA1* (P-value=2.20×10^-14^, OR=0.64 [0.58-0.72]). As discussed also previously by Degenhardt *et al.*^10^, *DQB1*\*06:02 is located on the same haplotype as *DRB1*\*15:01 (**Figure 2**). In our dataset, HLA-*DRB1*\*15 is the most significantly associated 1-field allele (OR=1.61 [1.40-1.85], P-value=2.07×10^-11^). Overall, the observed association of HLA alleles is in line with previous studies (**Figure 2**).

**Figure 2:**
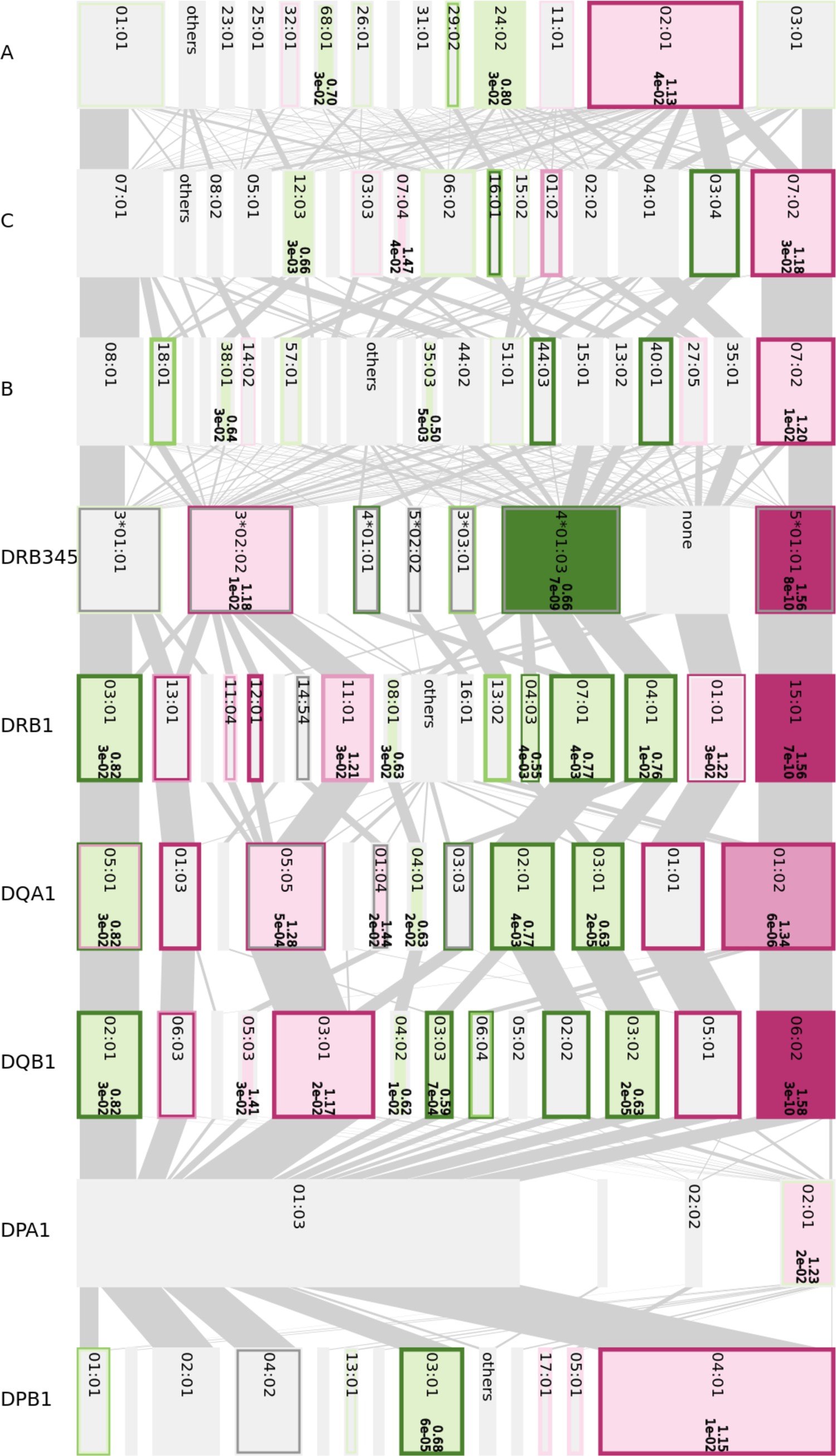
Disentangler42 plot that summarizes the HLA haplotype structure in our UC cases and controls. Risk alleles are colored in red (10^-5^<P-value<=0.05: light-red; 5×10^-8^<= P-value<10^-5^: mid-red; P-value<=5×10^-8:^ dark-red), protective alleles in green (10^-5^<P-value<=0.05 light-green; 5×10^-8^<= P-value<10^-5^: mid-green; P-value<=5×10^-8:^ dark-green), dark-grey missing data, light-gray alleles with no effect (P-value>0.05). The inner color of each box is based on our dataset, the inner frame of each box represents the results from Goyette et al.^9^ and the outer frame represents the European data from Degenhardt *et al.*^10^. (Note: We suspect that Goyette *et al.* did not distinguish between *DQA1*\*05:01 and *DQA1*\*05:05.) For genome-wide-significant alleles the OR and P-value is noted next to the allele names in bold type. The height of the box illustrates the allele frequency. Our results are concordant with the previous analyses by Degenhardt *et al.* and Goyette *et al.* as colors for each box are in most instances the same.

The main associated signal described by Goyette *et al.*^9^ HLA-*DRB1*\*01:03 is not nominally significant (cutoff P-value<1×10^-5^) in our data set due its low frequency in our data, even though its effect size is even larger in our German cohort (OR=5.31 [1.85-15.25], P-value=1.95×10^-3^, MAF_cases_=0.41%, MAF_controls_=0.96%) than reported by Goyette *et al.*^9^.

Of the 58 alleles previously reported to have a nominally significant association with UC either within the European dataset from Degenhardt *et al.*^10^ or the results from Goyette *et al.*^9^ only 6 showed an opposite direction of effect in our data. (**Supplementary Table 4** and **Figure 2** and **Supplementary Figure 32**). *DQA1*\*05:01 showed a risk effect in Goyette *et al.*^9^. However, we suspect that Goyette *et al.* named *DQA1*\*05:05 as *DQA1*\*05:01 since both belong to the same g-group of alleles and since the allele *DQA1*\*05:05 is not reported by Goyette, and the allele frequencies of *DQA1*\*05:05 (MAF=14.7%) and *DQA1*\*05:01 (MAF=11.9%) combined are closer to the allele frequency reported by Goyette *et al.* (MAF_cases_=28.0%, MAF_controls_=25.3%). Further, the associations of the alleles *A*\*29:02, *C*\*01:02, *DRB1*\*13:01, *DQA1*\*01:03 and *DQB1*\*06:03 were not supported in our dataset since all of them have an allele frequency below 10% in all cohorts and the lowest P-value in our data is with 0.28 for HLA-*DRB1*\*13:01 far from significant. Moreover, the three class II genes are located on the same haplotype (**Figure 2**).

As different alleles share attributes with each other on the protein level based on the amino acid sequence, we further analyzed the data for specificities in the amino acid sequence (**Supplementary Table 4**). Glutamic acid (D) at position 175 of DQA1 is the associated amino acid with the lowest P-value of 3.92×10^-12^ and an OR of 0.64 [0.56-0.72], the amino acid is only present in alleles with a protective direction of effect (e.g., *DQA1*\*03:01, *DQA1*\*02:01 and *DQA1*\*03:02). Alternatively, a Lysine (L) can be present at amino acid position 175, present mainly in the *DQA1*\*05 alleles, with different directions of effect or glutamine (Q) present mainly in the risk associated alleles and therefore also nominal significant associated (P-value of 3.92×10^-12^ and an OR of 0.64 [0.56-0.72]). The variation is based on rs2308891 (chr6:32642232). The amino acid is in the α2-domain of the protein.

Additionally, nucleotides and amino acids are stronger associated as the single alleles in our data set. The multiallelic variant rs9269955 (chr6:32584361) influences the amino acid 11 of the *DRB1* allele with the nucleotide G as the strongest associated (P-value of 5.08×10^-12^ and an OR of 1.50 [1.33-1.68], MAF=0.2920). Depending on the rs17878703 (chr6:32584360) characteristic either an allele of the *DRB1*\*15 and *DRB1*\*16 group with a proline (P) in position 11 (P-value of 1.99×10^-10^ and an OR of 1.54 [1.35-1.76]) or the *DRB1*\*01 group is present for this risk variant. This amino acid is involved in the interaction with the peptide in binding pocket 6 (peptide-HLA interaction as previously identified in Degenhardt *et al.* 2021^10^).

Other nucleotides and amino acids playing an important role in HLA-peptide interaction with genome-wide significant UC association are at amino acid positions 13 and 71 of the DRB1 protein, and amino acid position 86 of DQB1. All genome-wide significantly associated amino acids and nucleotides are listed in **Supplementary Table 4**.

### Binding motifs of associated HLA alleles

To figure out the similarity between the different UC-associated HLA alleles we generated a dendrogram based on the predicted binding peptides. The dendrogram of the HLA-DR-peptide interaction shows three groups of protective and risk alleles each (**Supplementary Figure 32**). The biggest cluster is the HLA-*DRB1*\*04 group with the main important alleles *DRB1*\*04:01 and *DRB1*\*04:03 but also all other *DRB1*\*04 alleles have at least a tendency to be protective if reported and except *DRB1*\*04:02 all cluster closely together. The Second protective cluster combines *DRB1*\*07:01 and *DRB1*\*09:01. Both are the only representatives of their 1-field allele group in all three datasets shown. *DRB1*\*04, *DRB1*\*07 and *DRB1*\*09 are the only alleles occurring together with the pseudogenes *DRB7* and *DRB8* and in most cases the protein coding *DRB4* gene. Therefore, this signal might be either based on the similarity between the alleles as all of them evolved most probably from the same ancestor^66^ or because of another locus that is in LD, e.g., especially the *DRB4* locus. The third protective cluster includes all present *DRB1*\*03 alleles (*DRB1*\*03:01 and *DRB1*\*03:02).

The risk alleles also cluster based on the 1-field classification in a cluster with *DRB1*\*01 (present as *DRB1*\*01:01 and *01:03) and *DRB1*\*15 (present as *DRB1*\*15:01, *15:02, *15:03 and only single cases of *15:06), additional *DRB1*\*12:01 was identified as risk allele but in this case the only other allele of the same serogroup *DRB1*\*12:02 does not support the same direction of effect but the allele is only present in very low frequencies and so far only Goyette *et al.* reached an association P-value below 0.05 for this allele.

For a closer look into the binding specificities, we generated logos for the genome-wide associated alleles (**Figure 3**). The separate clusters are characterized by individual binding characteristics. *DRB1*\*03:01 is especially characterized by an enrichment of acidic amino acids in binding pocket 4 and basic anchors in pocket 6 and 9. The *DRB1*\*04 alleles *DRB1*\*04:01, *04:03, and *04:04 are characterized by an antigen with a polar or acidic amino acid in position 6 with a residue with one or two carbon atoms. Similar characteristics are also present in anchor position 9 but here the size restriction is not as stringent and additional alanine (A) as a hydrophobic amino acid is present. *DRB1*\*07:01 and *DRB1*\*09:01 do not show any enrichment of acidic amino acids but in comparison to the risk alleles (**Supplementary Fig. 33**) they show an enrichment for the polar amino acids serine (S) and threonine (T) in position 4.

**Figure 3:**
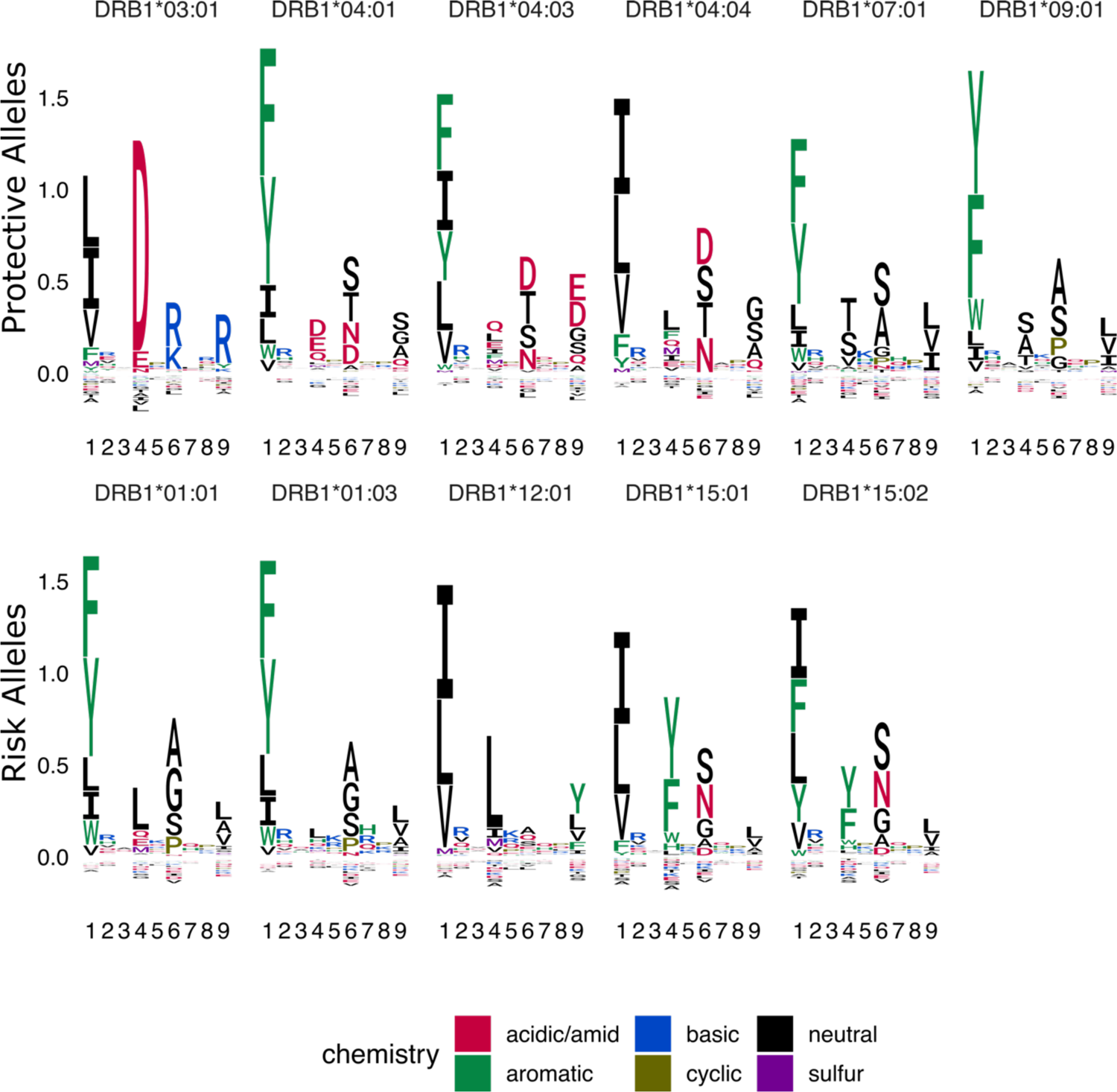
Binding logo plot of associated HLA-DR alleles. The upper row represents the protective associated alleles, the bottom line the logos of the risk alleles. The motifs are based on the NetMHCIIpan-4.0 predictions of the binding cores of all peptides at least annotated as weak binders. The single letters represent the one letter amino acid code colored by the chemical properties of the amino acids.

The risk alleles *DRB1*\*01:01 and *DRB1*\*01:03 are characterized by a small residue in binding pocket 6 (alanine, glycine, and serine) this can be explained by the high-volume amino acids in position 11 (L) and 13 (F) of the beta chain. *DRB1*\*12:01 peptides are characterized by a hydrophobic amino acid in pocket 4 (leucine or isoleucine) and a big uncharged amino acid in pocket 9 (tyrosine, phenylalanine, leucine, valine), one factor for this characteristic is that the allele has the smallest residue at position 9 (E) and smaller residues at position 39 and 57. The alleles *DRB1*\*15:01 and *DRB1*\*15:02 are mainly characterized by preferring aromatic amino acids in pocket 4 (tyrosine, phenylalanine, tryptophan).

Overall, the logo of the *DQ* alleles (**Supplementary Figure 34**) are not as well defined as the motifs of the *DR* alleles. Here, an acidic anchor occurs in position 6 of the risk *DQ* haplotypes *DQA1*\*01:01-*DQB1*\*05:01 and *DQA1*\*01:02-*DQB1*\*05:01 and an additional aromatic anchor in position 4 is present. Further, proline plays a more important role in binding peptides, but not in a disease-associated manner.

In summary, a single consistent binding motif for all risk or protective associated alleles cannot be defined. However, the protective alleles do tend to bind more often acidic amino acids, while the risk alleles are characterized by aromatic amino acids based on the binding predictions.

### PepWAS analysis

We performed a PepWAS analysis to investigate which self-peptides are differentially recognized by the UC risk and protective HLA-DRB1 proteins. For computational reasons we limited the analysis to UC susceptibility genes from previous studies and identified within this study testing the hypothesis that chronic inflammation in UC patients is driven by self-peptides encoded by the patient’s genome and that are presented by their own HLA molecules. Coding mutations in the genes encoding the peptides may additionally lead to the presence or absence of specific peptides in some patients and controls.

The PepWAS resulted in 234 significant peptides after multiple hypothesis correction (P-value<3.73*×*10^-6^ based on 13,411 peptides) for HLA-*DRB1* (**Supplementary Figure 35**, **Supplementary Table 5**). Those peptides originate from 46 of the 76 considered GWAS genes (in 155 transcripts). All peptides are specific to one of the genes. Up to 18 peptides identified through the PepWAS are annotated to one single gene. The number of peptides identified within a protein correlates with the length of the amino acid sequence (Pearson’s product-moment correlation: R^2^=0.27, P-value=1.02×10^-6^), especially when considering 2 peptides separately, if they have no overlap of 9 amino acids when shifted against each other (Pearson’s product-moment correlation: R^2^=0.44, P-value=4.71×10^-11^).

Eight of the proteins containing PepWAS hits are described in the meta-analysis from Linggi *et al.*^50^ or in the analysis of treatment naïve patients by Taman *et al.*^51^ as differentially expressed in ulcerative colitis patients. Whereof *JAK2*, *KIF21B*, *FCGR2A*, *NOD2,* and the HLA genes *DRB1* and *DQA1* are upregulated and *NXPE1* and *HSD11B2* are downregulated (**Supplementary Table 5**). Of those genes *KIF21B* and *HSD11B2* are neither reported as membrane proteins, nor as extracellular. Peptides of the genes *JAK2, FCGR2A* and *HLA-DRB1* were extracted with the peptidomes described in ElAbd *et al.*^53^. The only hit from the PepWAS analysis having at least nine amino acids overlap to the proteomes is the peptide RRVQPKVTVYPSKTS (P-value=1.52×10^-6^). This peptide is present in the sequence of alleles of the HLA-*DRB1*\*15, -DRB1*16, and -*DRB1*\*10 groups and of HLA-DRB4 alleles. The peptide was identified previously in 5 of 6 samples with a *DRB1*\*15:01 allele using antibody-based HLA-pulldown and LC-MS analysis^53^. The other sample has a genotype with two *DRB4* alleles, regarding the prediction the genotype of this patient (*DRB1*\*07:01/*DRB1*\*09:01) would not present this peptide. In the 9 other samples with a genotype with a DRB4 molecule but no *DRB1*\*15 allele (one case with 2 haplotypes with *DRB4*), the peptide was not found. Side note: Interestingly, all haplotypes with a copy of the *DRB4* gene (*DRB1*\*04, *DRB1*\*07 and *DRB1*\*09) are associated with a protective effect. 62 mutations are essential to generate one or more of the identified peptides, but thereof 40 would prevent the generation of another identified peptide. In comparison to the 22 missense mutations that lead to the generation of a peptide identified as associated in the PepWAS analysis, there are 156 missense mutations that lead to a peptide not being identified as significant in the PepWAS.

All PepWAS hits were predicted to bind HLA-DRB1*15:01 and the rare HLA-DRB1*15:06 protein. In most cases also the other tested DRB1*15 alleles (DRB1*15:02 and DRB1*15:03) were predicted to bind. Other alleles binding several hits are DRB1*16:01 and DRB1*16:02, the other alleles of the DR2 serotype and DRB1*01:03. Interestingly no peptide identified in the PepWAS was predicted to bind with DRB1*01:01, even though the allele shows a trend of being a risk factor.

The peptide hits are enriched for isoleucine, asparagine, valine, and tyrosine and have lower frequencies of cysteine, glutamic acid, glycine, and arginine compared to our candidate transcripts.

121 of the peptide hits were present in all individuals within our sample set, 133 are peptides of the reference proteome, of which 19 are not present in all individuals. The remaining 101 peptides require at least one mutation from the reference genome to be encoded (**Supplementary Table 5**). None of the mutations shaping the peptides was statistically significant associated with the disease after correcting for multiple testing and no peptide identified in the PepWAS analysis showed significantly different frequencies in cases and controls.

## Discussion

Different hypotheses on the underlying processes leading to inflammation in UC including the role of the HLA haplotype are discussed in the literature^8^. Here, we focused on the autoimmune hypothesis in association with autoantigens, excluding microbial peptide candidates from the analysis, because of the excessively larger search space. However, the binding preferences of the HLA alleles are general attributes and therefore parts of our results (and the analysis concept in general) can also be transferred to microbial candidates. While most of the identified genetic associations are not related to HLA antigen recognition, the consistent and strong association of HLA suggests that HLA peptide interaction plays an important role.

Our GWAS results are overall in line with previous studies. The previously not described genome-wide significant hit at 5p14.3 is either specific for our German subpopulation or represents a false positive signal. This intergenic association is well supported by variants in LD but could not be validated in the larger IIBDGC dataset. In addition, no biologically or disease relevant conclusions could be made.

The *NOD2* gene harbors the most prominent genetic associations known for CD. It has also been studied previously in the context of UC^62^, however, no clear associations for UC have been described before^7^. The herein determined novel association of rs61736932 with UC is supported by two large publicly available data sets^30,63^. Interestingly, the identified *NOD2* UC variants are in another location of the protein not in LD to those variants described for CD, suggesting a different functional role in UC.

The suggestively protective variant we identified in the *RBM19* gene, rs3782449, and the previously described risk variant with an association in ocular manifestation in IBD rs4766697^55^, are located on different haplotypes. The direction of effect for rs4766697 in our data is protective as well, even though far from statistical significance. To our knowledge, there are no other studies linking *RBM19* to IBD, therefore additional studies are needed to clarify the exact role of this candidate gene in UC disease etiology.

Previous GWAS already yielded over 240 genetic variants significantly associated with IBD^67^, but still those findings only cover a part of the heritability^68^. Our study cohort was not sufficiently powered to identify low-frequency variants and variants with small effect sizes. However, compared to previous GWAS, we used the genome build GRCh38 (others mainly used build GRCh37) and we employed state-of-the-art imputation reference and exome data sets. This enabled us to analyze previously not, or not properly, covered genetic regions. For example, the signal at 9q22.2 (*UNQ6494*) was not covered in GRCh37, which may explain why no former study identified this locus as associated with IBD. Nevertheless, an independent replication is necessary to validate this signal. Furthermore, for the bitter taste receptor gene *TAS2R43*, where we identified a suggestive hit without LD support in the exome data, the genetic architecture changed between the genome builds GRCh37 and GRCh38 and is still not well covered by imputation. This explains why a potential identification of disease-associated markers at this locus was nearly impossible beforehand but might be improved by further analyses of diverse whole genome sequencing data. It further shows the benefit of improving imputation reference panels and methods.

We identified two suggestive hits located within genes involved in the TNF pathway (*TNFRSF8* and *CTCF)*. TNF levels are typically increased in IBD patients and anti-TNF is a common treatment option for IBD even though the biological role of TNF is much more complex and treatment not effective in all patients^69^. Both signals were not validated in publicly available data sets.

The genetic risk profile of the HLA region in our data is consistent with UC associations described in the literature for Caucasian populations^9^. As shown by Degenhardt *et al.*^10^, this also holds true for different ethnicities in UC, even though some HLA alleles are not observed in Caucasian populations or more frequent in the non-Caucasian population.

The main genetic association signal for UC is located within the HLA-*DR* and HLA-*DQ* genes. Causality of either locus for UC cannot be shown with our data due to the strong LD between the loci. Mechanistic follow-up studies are clearly needed to disentangle this signal. On the computational level, Goyette *et al.*^9^ used a gene-based analysis and identified HLA-*DR* as the most likely disease-relevant candidate gene. However, as shown in **Figure 2**, and discussed in Degenhardt *et al.*, HLA-*DQ* cannot be fully ruled out, due to the very strong linkage disequilibrium with HLA-*DRB1*.

The PepWAS analysis ranks peptides based on the HLA profile of the disease-associated HLA alleles and enables the identification of disease-relevant binding motifs. However, the PepWAS approach cannot differentiate disease-relevant peptides from other similar peptides without a prefiltering of peptides of interest, due to peptide similarity among different proteins, statistical restrictions, and the limitation in the specificity of the HLA profile. In our analysis, 46 of the 76 GWAS candidate genes contained PepWAS hits. Longer protein sequences lead to a higher chance of identifying PepWAS hits within this sequence. This influence factor might be increased by the in-silico approach of defining the peptides by a sliding window. All candidates identified by PepWAS are DRB1*15:01 binders, this is associated to the fact that the allele has the largest power due to its frequency and comparably strong effect size. Alternative peptides would need to be predicted as binders for more than one serotype with the same direction of effect to be significant in the analysis.

The only peptide identified by PepWAS that was also present in the 25 immunopeptidomes described in ElAbd *et al.*^53^ is a peptide present in DRB1*15. It needs to be considered that the analysis of the immunopeptidomes was based on a reference dataset including only one sequence for each gene, and in case of *DRB1,* the sequence of this gene was DRB1*15:03. But the peptide in the peptidome is the only PepWAS hit in *HLA-DRB1*. The PepWAS was based on the personalized peptidomes of all patients and therefore included sequences of the different alleles in the dataset. Whether this peptide is a strong candidate to play a role in the pathogenesis of UC remains to be elucidated. On the one hand, this peptide is presented especially strong by DRB1*15 proteins, which are related to a higher risk. On the other hand, the peptide is present in the sequence of the protective haplotypes including a *DRB4* allele, and as the peptidomics data show that the peptide can be presented when carrying those haplotypes, even though reduced in comparison to individuals carrying a DRB1*15 allele. If the presentation of this peptide would play a significant role in modulating the disease, a strong protective effect would be expected e.g., for DRB1*08 alleles, where the peptide is not strongly bound nor present in this haplotype. However, such an effect could not be observed.

The peptide sets predicted to bind the risk alleles are characterized by aromatic amino acids in pocket 4, while the peptides binding the protective alleles are characterized by acidic amino acids in pocket 4. However, both characteristics do not apply to all the significantly associated alleles and no peptide identified in the PepWAS analysis is encoded with significantly different frequencies between our UC patient and control exome data.

Peptides containing mutation sites are less likely to be identified by our PepWAS analysis. One explanation for this might be a bias based on the training dataset used for the prediction algorithm. As the peptidome used for NetMHCIIpan-4.0^46^ training was derived mainly from mass-spectrometry data using the human reference proteome to define the peptides, and while the negative peptides were defined by sampling from the UniProt database, the binding peptides in the prediction might be biased towards the human reference proteome. Besides, the version 4.0 of NetMHCIIpan^46^ is based on immunopeptidome data and therefore the training data is expected to be closer to the *in vivo* situation than the previous versions that were based on peptide competition assays. Still, the prediction of HLA-peptide interaction does not cover the T cell specificity and therefore presents only a necessary but not complete part of an HLA-induced immune reaction.

In summary, we employed a large exome dataset from UC patients and newly available reference datasets to obtain further insights into the role of the HLA in UC disease etiology. The initial association analysis identified promising genetic signals in *NOD2*, *RBM19*, *TAS2R43*, as well as at the intergenic loci 5p14.3 and 9q22.2. Further, additional suggestive hits related to the TNF pathway were identified. An additional replication of those associations is highly recommended but as most of them either have relatively low frequencies or are not well covered in older genome references, a replication within a large new dataset is necessary. A gene-based analysis on the rare variants revealed by exome-sequencing did not result in any significant results, but as for the three genes with the lowest P-value, an already described connection to the disease could be drawn, it is expected that a more powerful analysis including more samples would result in significant and relevant findings.

Our analysis of the HLA showed again a stable association of UC with multiple HLA alleles. Here, we were also able to highlight some characteristics of the peptides interacting with the HLA even though no specific autoimmune peptide candidates could be identified, where a mutation impacts the peptidome in a disease-specific way. This supports either the hypothesis of an external/environmental origin of a pathogenic peptide, for example from the microbiome or the diet, or an autoimmune interaction independent from non-synonymous mutations.

## Materials and Methods

### Cohort description

In total 15,877 individuals from studies across Northern Germany, with mainly chronic inflammatory traits (arthritic dermatitis, Crohn’s disease (CD), longevity, primary sclerosing cholangitis, psoriasis, sarcoidosis, and ulcerative colitis (UC)) and 4,680 population controls with unknown phenotypes were genotyped and submitted to joint genotyping quality control as described in the section *Array-based genotyping, quality control and imputation* as a resource for the genetic analysis of inflammatory diseases. For reasons of feasibility, this study focuses on the analysis of UC only, since UC exhibits the strongest HLA associations. For this purpose, we included 863 ulcerative colitis patients and 4,185 population controls from the total available quality-controlled genotype cohort. The population control is comprised of 969 individuals recruited within the German Food Chain Plus (FoCus) cohort, previously described in Barbaresko *et al.*, 2020^12^ and 3,216 German healthy blood donors as used and described in Degenhardt *et al.* 2022^13^. Of the 863 UC patient samples 424 were previously described by Franke *et al.*, 2008^4^ and 439 by Bokemeyer *et al.,* 2016^14^.

In total 5,048 individuals (4,185 controls, 863 UC patients) were used for analysis. A detailed overview of sample numbers is shown in **Supplementary Table 1**.

### Ethics approval

The study was conducted according to the guidelines laid down in the Declaration of Helsinki and was approved by the Ethic Committee of the Medical Faculty of the University of Kiel (Germany). All participants gave written informed consent, and the recruitment protocols were approved by the ethics committees at the respective recruiting institutions. The following approvals of the project were obtained from the ethics committees: BioColitis samples (D 474/12)^14^, German blood donors (A 103/14)^13^ and the samples of the FoCus Cohort as well as the samples described in Franke *et al.*, 2008^4^ (A 156/03).

### Sample preparation/processing

DNA was extracted by the DNA laboratory of the Institute of Clinical Molecular Biology (Kiel University, Kiel, Germany) from whole blood. For a detailed description of the further extraction protocol, we refer to Ellinghaus *et al.*^15^.

### Array-based genotyping, quality control and imputation

Genotyping was performed using the Global Screening Array (GSA), version 1.0, containing 700,078 variants pre-quality control at the Regeneron Genetics Center, U.S.A. The data were called on genome build GRCh37 (using the cluster file GSAMD-24v1-0-A_4349HNR_Samples.egt). Genotype quality control (QC) was performed as implemented by BigWAS^16^. Briefly, the variants are annotated in a standard way based on a database and filtered on missingness (≥ 0.02 in a single batch or ≥ 0.10 in all batches), and the Hardy-Weinberg equilibrium (false discovery rate (FDR) threshold of 10^-5^ in controls). Then samples with high missingness (≥ 0.02), increased or decreased heterozygosity rates (± 5 standard deviation), relatedness testing (identity by descent ≥ 0.1875) and based on the population structure identified by principal component analysis (PCA) (outside the ± 5*IQR in PC1 and PC2) are removed. Of the final sample set, additional variants are filtered for differential missingness between controls and diseased samples and monomorphic sites.^16^ After QC, a total of 5,048 individuals (863 cases/ 4,185 controls) and 579,352 variants remained for analysis (**Supplementary Table 1**). Only variants mapping to the autosomes were used here for association analysis. All genomic positions were lifted to genome build GRCh38 for further analysis on the TOPMed Imputation Server or for the genotyped GSA data within the BigWAS^16^ pipeline using the UCSC liftOver tool^17^. To increase the genotyping coverage, SNP imputation was performed using the TOPMed Imputation Server from the NIH using the TOPMed Imputation diverse reference panel (version TOPMed-r2@1.0.0) including 97,256 deeply sequenced human genomes with a post-imputation quality score of R^2^ set to 0.1^18–20^. In total 90,406,930 variants had an R^2^ larger than or equal to 0.1. In the following analysis 13,780,246 variants with an R^2^ > 0.6 and a MAF above 1% were analyzed if not noted otherwise.

### Exome sequencing, genotyping and quality control

Whole exome sequencing (WES) was performed at the Regeneron Genetics Center, U.S.A. Sample preparation and sequencing as well as the sequence alignment, variant identification and genotype assignment were done as described in Van Hout *et al.* 2020^21^. An extended quality control was conducted with the ‘Goldilocks’ (GL) filters^21^ and additional filters for genotypes, variants and samples. In brief genotypes were set to “no call” based on the WeCall filters allele bias (ABPV <0.009), allele and strand bias (ABPV + SBPV <0.07), bad reads (BR <15), low quality (LQ <10), low mapping quality (MQ <40), quality over depth (QD <15) and strand bias (SBPV <0.01) and further, based on the sequencing depth (DP <7), genotyping quality (GQ >14) or allele balance (AB <0.25). Variants were removed if no reliable sample (AB ≥0.15 or homozygous) remained. For insertions and deletions (InDel) the same filtering was used with different parameters: genotypes with a DP <10 were removed and a reliable sample to keep a variant was defined by an AB ≥0.20 or homozygosity.^21^ Further, single nucleotide variants (SNVs) were removed if the missingness was above 10% (variants that overlapped with an InDel were not considered for missingness filtering) or if the Hardy-Weinberg-Equilibrium P-value in control individuals was below 10^-5^.

Individuals were removed if they presented with an unusual rate (> 6 standard deviation difference to the mean) of (a) singletons (more than 737) or (b) missingness (equaling a missingness cutoff of 0.21), or (c) heterozygous to homozygous ratio (0.0058 < hethom < 0.0077) or (d) transition/transversion ratio (2.14 < Ti/Tv < 2.43). Additionally, individuals were removed if the reported sex disagreed with the genetic sex. See **Supplementary Table 1** for the sample numbers removed by the different criteria.

Finally, 5,390,149 variants passed exome sequencing filters, summarized in 5,033,063 non-overlapping positions of variation. 19,688 individuals passed QC, of which 17,138 individuals overlapped with the quality-controlled genotyping dataset described above and were used for further analysis.

### Genome-wide association analysis

Genome-wide association tests were conducted using SAIGE^22^ (0.45.0) on both the variants from exome sequencing and genotype imputation implemented within the BIGWas pipeline that is available at GitHub ikmb/gwas-assoc^16^. In brief, a logistic mixed-effects model was applied on the UC case-control status using genotype dosage (imputed data) or genotype hard-call genotypes (exome data). Genotype dosages were used to appropriately consider imputation uncertainty. The model additionally included the first 10 PCs calculated from the quality-controlled genotypes from the GSA (pre-imputation and post-quality control). We calculated the genomic inflation factor (lamdaGC) with and without excluding the HLA region (chr6, 29Mb-34Mb)^23^. Variants with a P-value of association < 5×10^-8^ were defined as genome-wide associated with UC, while variants with a P-value of association < 10^-5^ were considered to have at least a nominal (suggestive) association. Bayesian fine mapping was performed using the tool FINEMAP (version 1.4) with the parameters --n-causal-snps 1 --sss (shotgun stochastic search)^24, 25^.

Linkage-Disequilibrium-based (LD-based) clumping was calculated using PLINK with a significance threshold for the index SNP (clump-p1) of 0.00001 and a secondary significance for clumped SNPs (clump-p2) of 0.001 in a range of 150 kilo base pairs (kbp).

Gene expression impacts for the associated variants was looked up using the R-package Qtlizer^26^ and gtex associated genes with a P-value below 10^-5^ were considered marginally associated.

### Replication of suggestive hits

All suggestive variants were investigated extensively using regional association plots created with LocusZoom^27, 28^. Variants with insufficient LD-support (less than two additional variants with P-value <10^-3^ in ±150 kbp) and a MAF < 1% were discarded as false positive associations. To validate all remaining hits, we performed a lookup of variants, considering ±150 kbp around our lead SNP in the NHGRI-EBI GWAS Catalog v1.0.2^29^ and in the Rapid Imputation for COnsortias PIpeLIne (RICOPILI; dataset IBD_UC_1KG_oct13 dataset^30^) considering variants with linkage-disequilibrium (LD) R^2^ > 0.9 from our lead-variants^30^. LD was calculated on the exome and TopMED imputed data using PLINK^31^. Variants were considered to replicate if the lead variant itself or variants in high LD (R^2^ > 0.9) had a P-value of association below 0.05.

Further, a power analysis based on the NHGRI-EBI GWAS Catalog of genome-wide association studies^29^ data was performed to investigate replicability of known IBD variants in our dataset at a P-value threshold of < 10^-5^. For this purpose, odds ratios (OR) reported in the GWAS Catalog were used, together with the sample size (n_cases_=863, n_controls_=4,185) and allele frequencies derived from this study’s data. As the power calculated by a single analysis tends to be overestimated (referred to also as the winner’s curse)^32–34^, we computed a corrected value for power as the median power for the respective reported lead variant within neighborhood of ±5,000 base pairs under exclusion of the highest power value if more than one study was reported in the NHGRI-EBI GWAS Catalog.

### Gene based analysis

To include the genetic variation based on rare variants (MAF < 1%), which have limited power in the classical analysis, a gene-based analysis was performed using SAIGE-GENE^35^. SAIGE-GENE performs three different tests: 1) The Burden test analyses the correlation between the number of variants and the disease status and is therefore powerful if the single variants show the same direction of effect. 2) The sequence-based kernel association test (SKAT) aggregates the test statistics of the single variants and is therefore robust against variations in the direction of effect. 3) The SKAT-O is a linear combination of the other two tests. For more details see Lee *et al.* 2012^36^ and Zhou *et al.* 2020^35^. The analysis focusses only on the exome sequencing data, as we are focusing here on the genes and as rare variants cannot be imputed in sufficient quality. All variants with an allele-frequency <1% are grouped based on the gene-annotations generated by bcftools/csq^37^ using release 100 of the primary assembly of the human proteome from Ensembl^38, 39^. A relatednessCutoff of 0.125 and the same 10 PCs as for the GWAS analysis were used. Additionally, the following parameters were applied: minMAC=0.5, LOCO=FALSE, IsSingleVarinGroupTest=TRUE, IsOutputAFinCaseCtrl=TRUE, IsOutputNinCaseCtrl=TRUE, IsOuputHetHomCountsinCaseCtrl=TRUE. A gene is considered genome-wide significant if the P-value is below 2.62×10^-6^ (based on 19,117 genes).

### HLA imputation and fine mapping analysis

Imputation of alleles in the HLA region was performed for the classical HLA class I loci HLA-*A*, -*B*, -*C* and the class II loci HLA-*DQA1*, -*DQB1, -DPA1, -DPB1,* -*DRB1, -DRB3/4/5* at 2-field full context resolution from quality-controlled SNP genotype data. Here, we extracted pre-imputation SNP genotypes from the extended HLA region (chromosome 6: 29-34Mb) and used them as input for HLA genotype prediction with the random-forest based machine learning tool HIBAG (version 1.20.0)^40^ using the multi-ethnic reference model published by Degenhardt *et al.*^41^, which was modified to include the variants available on the GSA. We additionally derived amino acid and additional SNP imputation and defined marginal posterior probabilities for single HLA alleles across all predicted HLA genotypes as described in Degenhardt *et al.*^41^. Classical HLA alleles with a marginal posterior probability from imputation <0.3 were further excluded. Association analyses were conducted using PLINK’s logistic regression framework on the UC case/control status using hard-call HLA alleles, SNPs and amino acids from the imputation and the first 10 PCs calculated on whole-genome genotype information pre-imputation and post-quality control. For a closer look into the HLA haplotype structure, the tool disentangler was used^42^.

### Generation of personalized proteomes

For subsequent downstream analysis of genome-wide SNP data, a personalized haplotype-aware consequence-caller was used to predict the effect of genetic variants observed in the study cohort at the protein level. These genetic variants were obtained by merging WES data with the quality-controlled genotype data from the GSA (pre-imputation) using the Ensembl human genome FASTA release 100^38, 39^ as a reference. Four main steps were performed: (1) Phasing of nucleotides from WES with eagle (v2.4.1)^43^, (2) calling of the variant effect at the protein level (*i.e.,* the type and if available the effect of a variant on the protein level, for example, a missense variant chr4:85742C>T in ENST00000609518 with the exchange 48P>48S), (3) filtering data for chosen candidate genes (4) translation into FASTA files. (1) To enable the phasing of the sparse WES data the variant files derived from WES and GSA-based genotyping were merged. Further, multiallelic variants were transformed into different biallelic variants with bcftools/norm^44^ as eagle is not capable of dealing with multiallelic variants. The phasing was then conducted using eagle (v2.4.1)^43^ without an external reference. The resulting file was scanned for contradictory phased variants and rectified randomly by changing the phase for one of the contradictorily phased genotypes. In detail, as multiallelic variants *e.g.,* ref/alt1/alt2 are split (ref/alt1 + ref/alt2) and phased independently, for the rare case of heterozygous alternative variants (genotype alt1/ alt2) the phasing might lead to the phased genotype ref|alt1 + ref|alt2 which is contradictory as the merging back of the overlapping variants would lead to the ref in one haplotype and the alt1 and alt2 both in the second haplotype. (2) The haplotype aware variant effects were predicted using bcftools/csq^37^ on the merged WES/GSA dataset resulting in a VCF file containing 1,353,644 protein changing variants (1,250,354 missense variants, 6,830 inframe-insertions, 17,909 inframe-deletions, 46 inframe-altering variants, 4,904 start-loss variants, 43,201 stop-gain variants, 2,652 stop-loss variants and 43,994 frameshifts^1^). (3) For the following parts the set of genes was filtered to those with an association to UC. Here, we considered genes described previously as associated with UC at a genome-wide significant level in at least 4 different UC studies integrated in the NHGRI-EBI GWAS Catalog or variants not described in the catalog but observed here to have a nominally significant association (for the genotyping data additionally LD support is mandatory) with UC in this study. This resulted in a selection of 76 genes with 322 protein coding transcripts. A list of these genes and transcripts is shown in **Supplementary Table 2**. (4) We utilized VCF2Prot^45^ to generate a per-patient personalized version of all proteins transcribed from these 322 candidate transcripts by including the amino acid changes of step (2) into the reference sequences from Ensembl release 100^38, 39^. Briefly, the VCFs from step (2) were filtered for records containing the target transcripts into a new VCF file. The newly generated files along with the reference protein sequence of each transcript were fed into VCF2Prot and the results were stored as a FASTA file per patient containing all mutated isoforms.

### Peptide binding prediction

The generated personalized protein sequences for the set of candidate transcripts, along with the reference protein sequence were fragmented into 15-mer peptides using a sliding window approach with a step size of 1 and the generated results were stored in an SQL database for downstream analysis.

From all records stored in the database, we next created a set of unique peptides across all transcripts and patients. Additionally, we created a set of unique HLA-*DR* alleles and HLA-*DQ* alleles observed across all patients from the HLA imputation described above. For all combinations of unique peptides and unique HLA alleles from these lists, HLA-peptide binding affinities were predicted using NetMHCIIpan-4.0^46^.

### Binding motifs

The predicted binding affinities were used to generate a sequence logo of the predicted bound peptide repertoire for the associated HLA-*DRB1* alleles and HLA*-DQ* (genome-wide significant (P <5×10^-8^) in Goyette *et al.*, the European dataset of Degenhardt *et al.* or our dataset and no other effect direction in one of the other studies, for *DQA1* and *DQB1* pairings these conditions need to be true for both genes independently). This representation visualizes an enrichment or depletion of amino acids in comparison to the background amino acid frequency. The peptides are truncated to the 9-mer core predicted by NetMHCIIpan.

We generated 2 logos for each allele: (1) The logo generated by including all peptides predicted as weak or strong binder (%rank score <10). (2) Only the peptides that additionally fulfill the criterion of not binding against any of the significant alleles with the opposite direction of effect. The logos present the probability-weighted Kullback-Leibler logos with pseudo counts^47^. The logos were adjusted with pseudo counts based on the BLOSUM 62^48^ substitution matrix using a ß of 200, therefore adding 200 artificial peptides reflecting typical evolutionary mutations^47^. The graphical presentation was then performed using the R package *ggseqlogo*^49^.

### Peptidome-wide association study (PepWAS)

The predicted peptide binding affinities were further used for a PepWAS analysis. Peptides with a percentile rank score below 2% (strong binder) were considered to be presented by an HLA protein. Finally, the set of bound peptides were combined with the patients’ phenotypes and HLA genotypes to conduct a PepWAS as described initially by Arora and colleagues^11^. The aim of PepWAS is to identify peptides that might be relevant for immune recognition based on their binding affinity. In brief, PepWAS discriminates peptides based on the predicted binding affinity in patient versus control samples. Therefore, the same logistic regression model as for the genetic association (GWAS) analysis was used including the first 10 PCs.

The proteins and their peptides are analyzed for supporting factors. Those are: (1) Mutations in our sample set influencing the presence of the single peptides. (2) The gene expression signature in ulcerative colitis patients as published by Linggi *et al.*^50^ and by Taman *et al.* ^51^. (3) The cellular compartment of the genes^52^. The information of the subcellular compartments was exported from https://download.jensenlab.org/human_compartment_knowledge_full.tsv on 25.10.2022, the highest score presented for the gene ontology term GO:0005886 (membrane proteins) and GO:0005576 (extracellular proteins) are considered relevant. (4) A comparison with immunopeptidome data previously published and described by ElAbd *et al.*^53^.

## Acknowledgements

-

## Contributions

M.W., A.F. conceived and initialized the project. M.W. and H.E. analyzed the data, with help of M.H., F.U.-W., E.M.W., L.W., S.J., R.G.C., and D.E.. M.W. wrote the manuscript with help of H.E.. M.L., M.Z., B.B., and S.S. collected the samples. R.G.C. generated the WES data. H.E., P.B., T.K., and A.T. generated the peptide elution data. T.L.L. and A.F. supervised the project.

All authors revised and edited the manuscript for critical content and approved of the final version to be published.

## Data Availability

All data produced in the present study are available upon reasonable request to the authors.

## Footnotes

1 The sum of mutations by types are more than the overall number, as the variants include multiallelic variants and mutations might have different effects in different transcripts.

