## Supplement for "Genome-wide analysis of individual coding variants and HLA-II-associated self-immunopeptidomes in ulcerative colitis"

##### Supplementary Figures:

**Supplementary Figure 9:** Regional association plot for the locus 5p14.3 with the top hit chr5:18748431 (rs2937516). . .......... 11
**Supplementary Figure 10:** Regional association plot for the locus 5p13.1 with the top hit chr5:40323836 (rs348594). . .......... 12
**Supplementary Figure 11:** Regional association plot for the locus 6p21.33 with the top hit chr6:31118325 (rs117198148). . .......... 13
**Supplementary Figure 12:** Regional association plot for the locus 6p21.32 with the top hit chr6:32644620 (rs6927022). . .......... 14
**Supplementary Figure 13:** Regional association plot for the locus 9q22.2 with the top hit chr9:89844860 (rs36147380). . .......... 15
**Supplementary Figure 14:** Regional association plot for the locus 10q24.2 with the top hit chr10:99541336 (rs4590800). . .......... 16
**Supplementary Figure 15:** Regional association plot for the locus 12p13.31 with the top hit chr12:9333053 (rs187033004). . .......... 17
**Supplementary Figure 16:** Regional association plot for the locus 12q24.13 with the top hit chr12:113880288 (rs3782449). . .......... 18
**Supplementary Figure 17:** Regional association plot for the locus 16q12.1 with the top hit chr16:50666737 (rs139397276). . .......... 19
**Supplementary Figure 18:** Regional association plot for the locus 16q22.1 with the top hit chr16:67493201 (rs77919558). . .......... 20
**Supplementary Figure 19:** Regional association plot for the locus 19q13.11 with the top hit chr19:32088213 (rs6510221). . .......... 21
**Supplementary Figure 20:** Regional association plot for the locus 19q13.31 with the top hit chr19:43804850 (rs364691). . .......... 22
**Supplementary Figure 21:** Regional association plot for the locus 22q13.1 with the top hit chr22:39318699 (rs1569498). . .......... 23

### Supplementary Figures

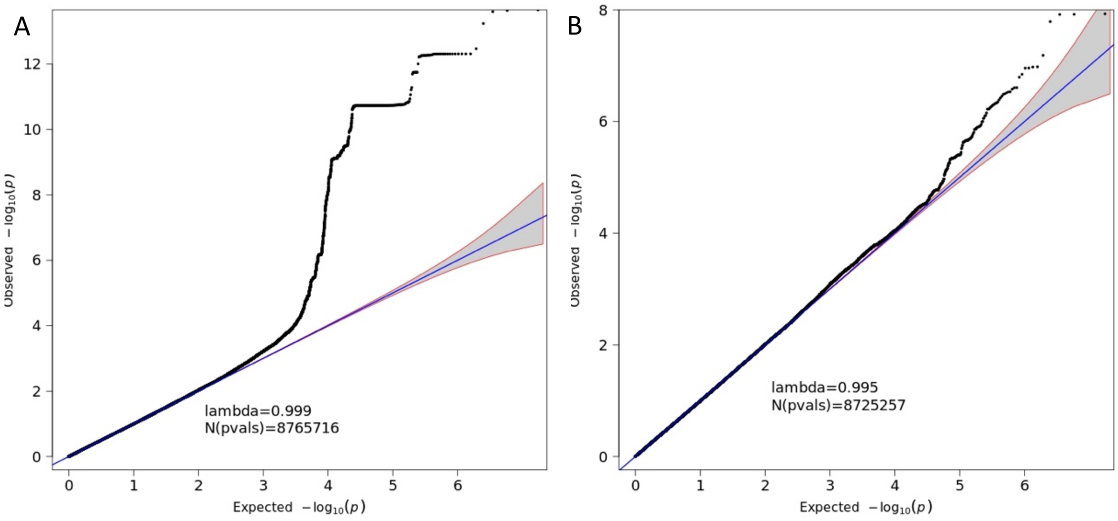

***Supplementary Figure 1:*** *Quantile-quantile plot of association summary statistics of the imputed genotyping data. The 95% concentration band under random sampling is shown in gray. The genomic inflation factor lambda is defined as the ratio of the medians of the sample χ2 test statistics and the 1-df χ2 distribution (0.455).*[1] *Panel* ***(A)*** *includes all 8,765,716 variants with MAF > 1 % and an imputation score* $r^2$*>0.6. Panel* ***(B)*** *excludes the variants of the HLA-region (chr6:29-34MB).*

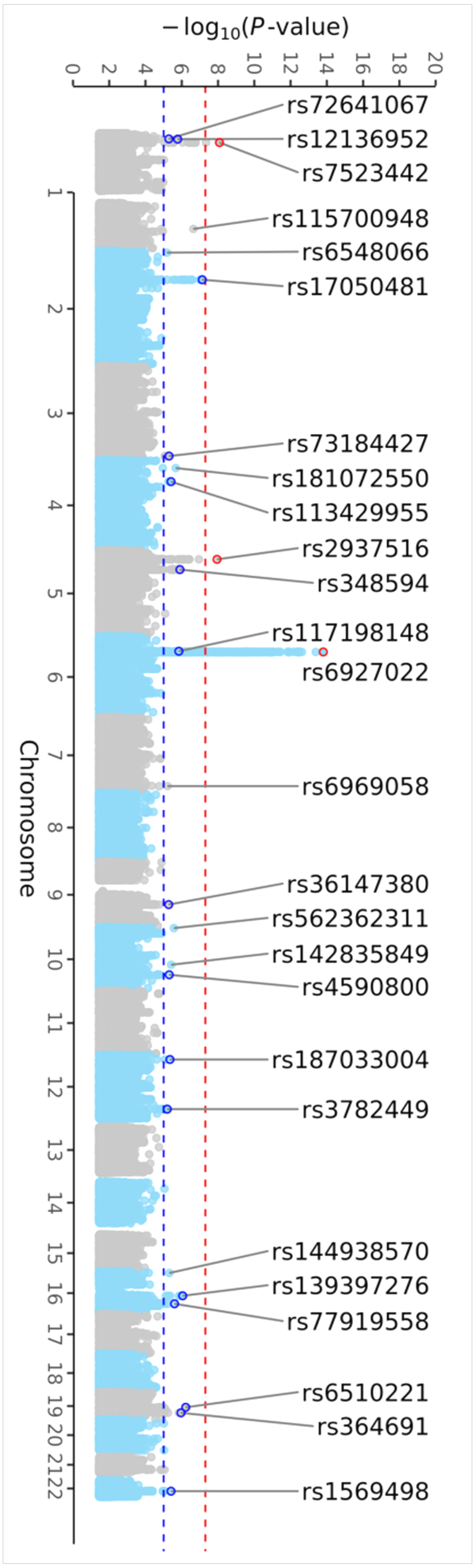

***Supplementary Figure 2:*** *Manhattan plot of the imputed genotyping data with a MAF > 1 % and an imputation score $r^2$>0.6. All loci of at least nominal significance (blue horizontal line; $P<1\times10^{-5}$) are annotated by the SNP-ID. Loci with LD support are highlighted with a blue (nominal significance) or red circle (genome-wide significance, red horizontal line; $P<5\times10^{8}$).*

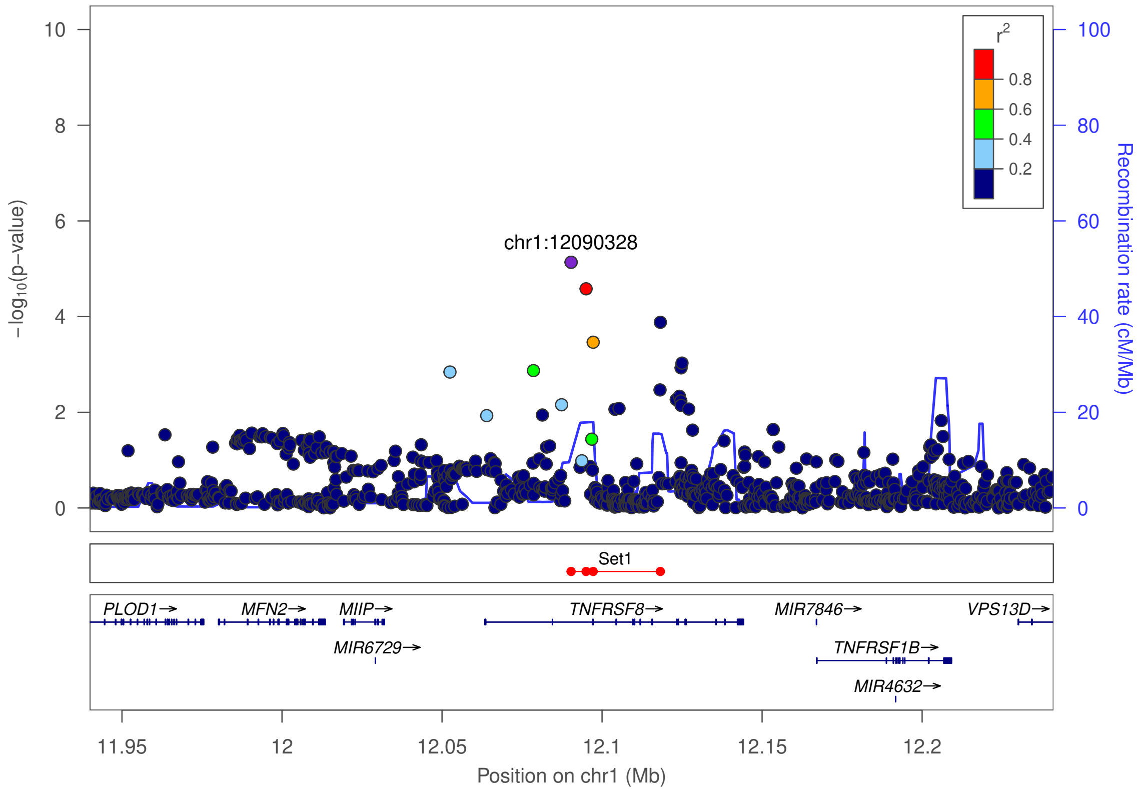

***Supplementary Figure 3:*** *Regional association plot for 1p36.22 with the top hit chr1:12090328 (rs72641067). The purple dot represents the most strongly associated SNP with ulcerative colitis. The color of the dots represents the linkage disequilibrium (LD) with the most strongly associated SNP (see color legend). The positions represent the genome build GRCh38. The recombination rate is shown in centimorgans (cM) per million base pairs (Mb). The bottom part shows the name and locations of the genes within the region. The thicker blue line represents the position of the exons, while the thinner line represents the intronic regions. The direction of transcription is represented by an arrow behind the name of the gene. The plot was created using LocusZoom*[2].

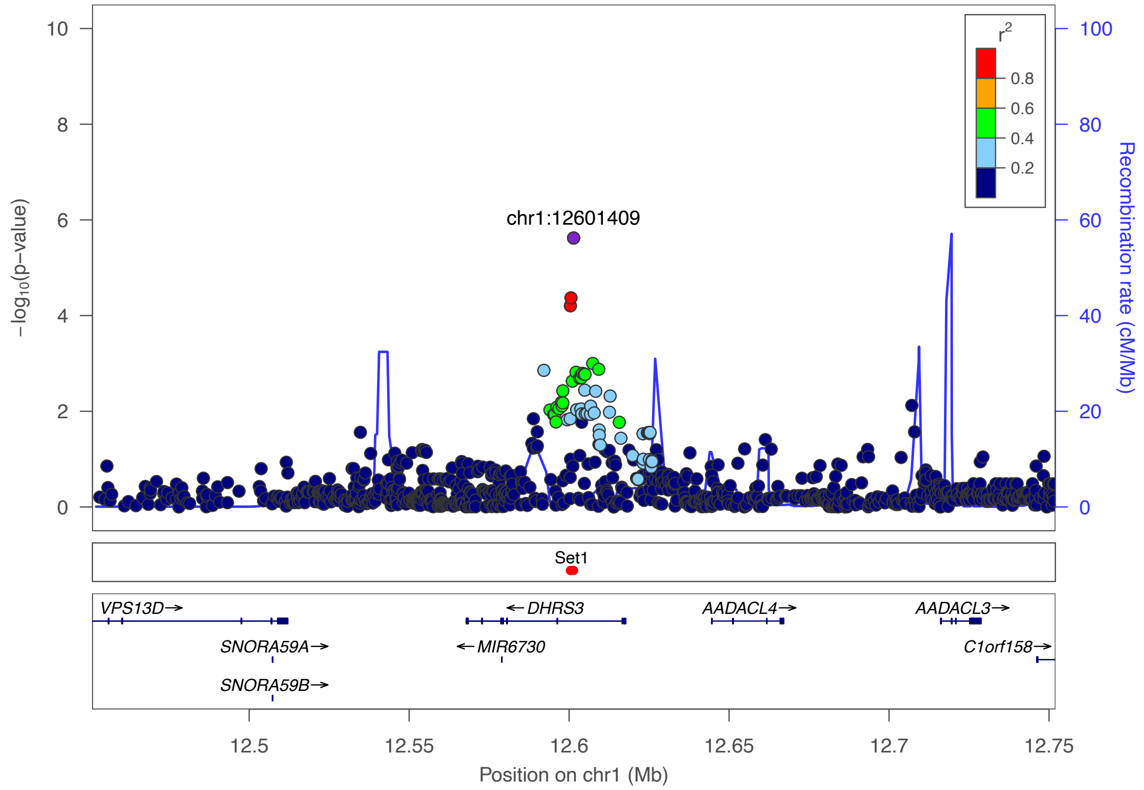

***Supplementary Figure 4:*** *Regional association plot for the locus 1p36.22 with the top hit chr1:12601409(rs12136952). The purple dot represents the most strongly associated SNP with ulcerative colitis. The color of the dots represents the linkage disequilibrium (LD) with the most strongly associated SNP (see color legend). The positions represent the genome build GRCh38. The recombination rate is shown in centimorgans (cM) per million base pairs (Mb). The bottom part shows the name and locations of the genes within the region. The thicker blue line represents the position of the exons, while the thinner line represents the intronic regions. The direction of transcription is represented by an arrow behind the name of the*

*gene. The plot was created using LocusZoom[2].*

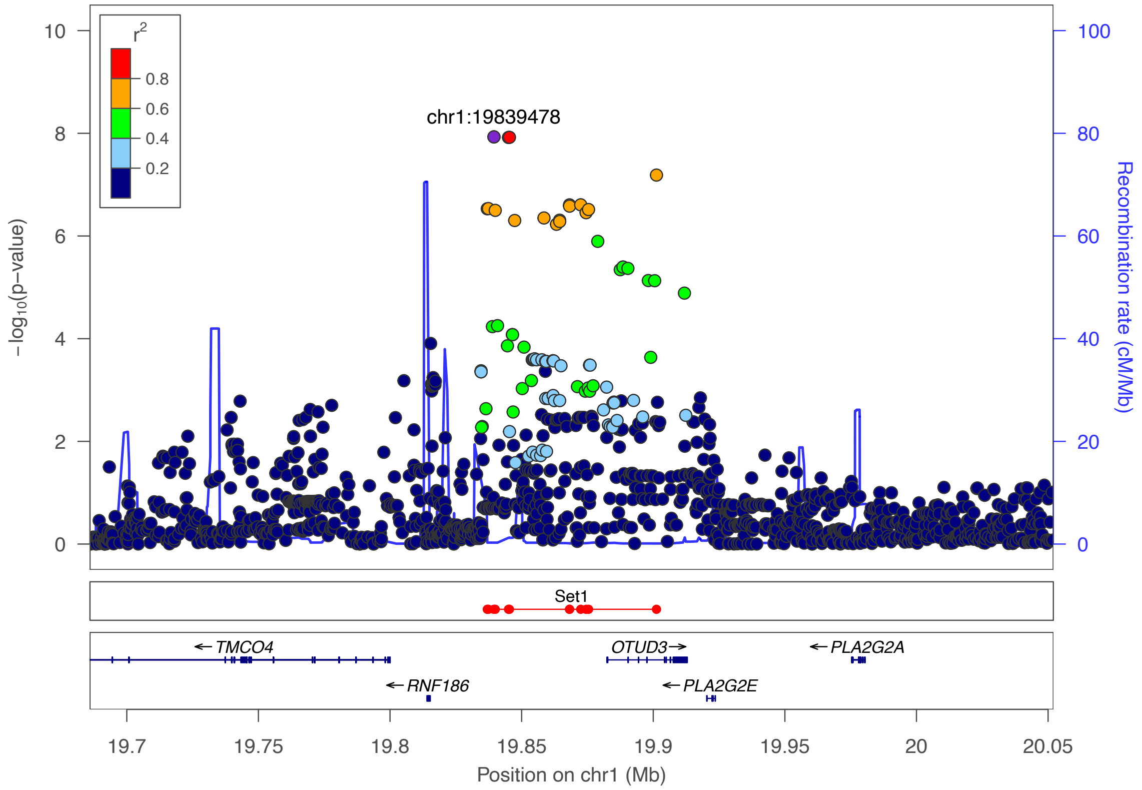

***Supplementary Figure 5:*** *Regional association plot for the locus 1p36.13 with the top hit chr1:19839478 (rs7523442). The purple dot represents the most strongly associated SNP with ulcerative colitis. The color of the dots represents the linkage disequilibrium (LD) with the most strongly associated SNP (see color legend). The positions represent the genome build GRCh38. The recombination rate is shown in centimorgans (cM) per million base pairs (Mb). The bottom part shows the name and locations of the genes within the region. The thicker blue line represents the position of the exons, while the thinner line represents the intronic regions. The direction of transcription is represented by an arrow behind the name of the gene. The plot was created using LocusZoom[2].*

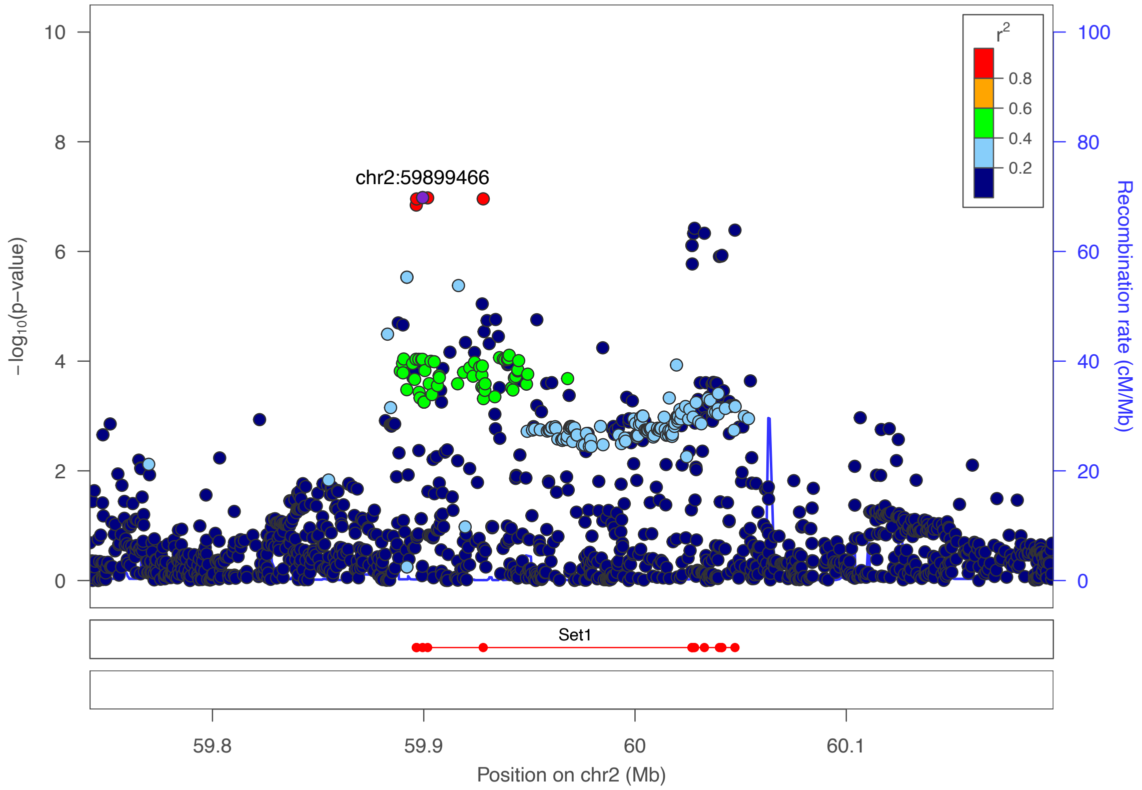

***Supplementary Figure 6:*** *Regional association plot for the locus 2p16.1 with the top hit chr2:59899466 (rs17050481). The purple dot represents the most strongly associated SNP with ulcerative colitis. The color of the dots represents the linkage disequilibrium (LD) with the most strongly associated SNP (see color legend). The positions represent the genome build GRCh38. The recombination rate is shown in centimorgans (cM) per million base pairs (Mb). The bottom part shows the name and locations of the genes within the region. The thicker blue line represents the position of the exons, while the thinner line represents the intronic regions. The direction of transcription is represented by an arrow behind the name of the gene. The plot was created using LocusZoom[2].*

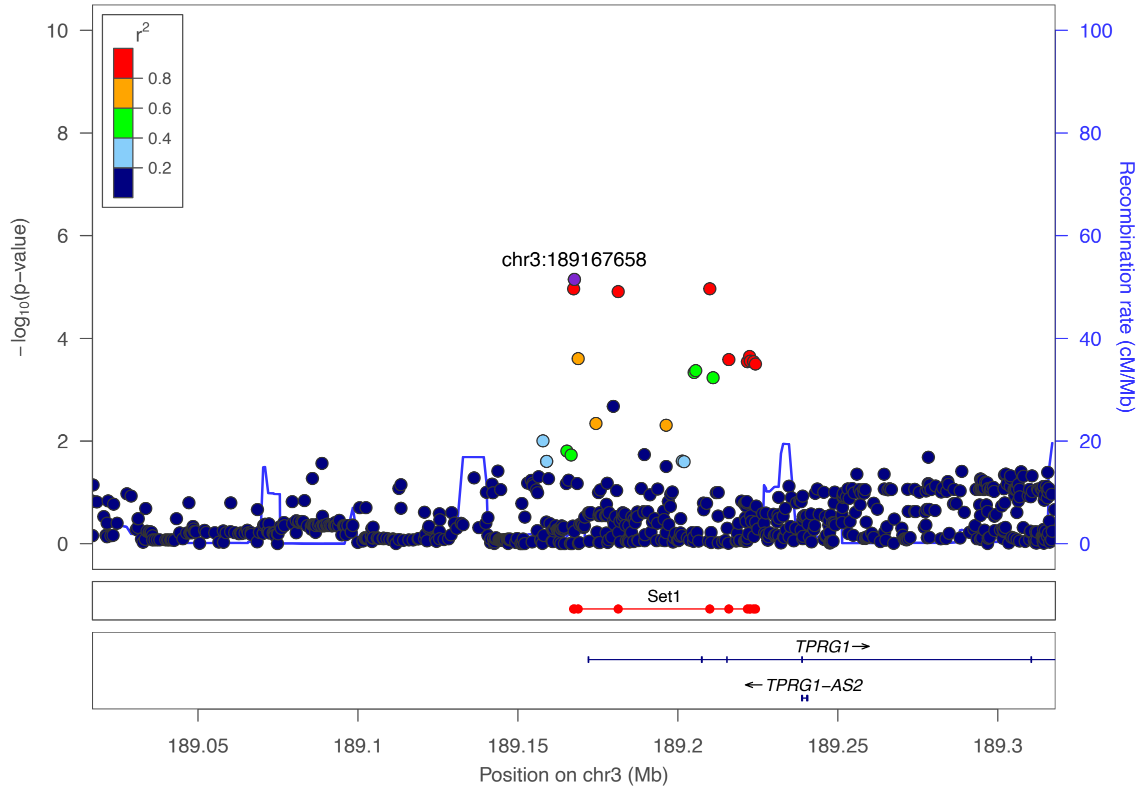

***Supplementary Figure 7:*** *Regional association plot for the locus 3q28 with the top hit chr3:189167658 (rs73184427). The purple dot represents the most strongly associated SNP with ulcerative colitis. The color of the dots represents the linkage disequilibrium (LD) with the most strongly associated SNP (see color legend). The positions represent the genome build GRCh38. The recombination rate is shown in centimorgans (cM) per million base pairs (Mb). The bottom part shows the name and locations of the genes within the region. The thicker blue line represents the position of the exons, while the thinner line represents the intronic regions. The direction of transcription is represented by an arrow behind the name of the gene. The plot was created using LocusZoom[2].*

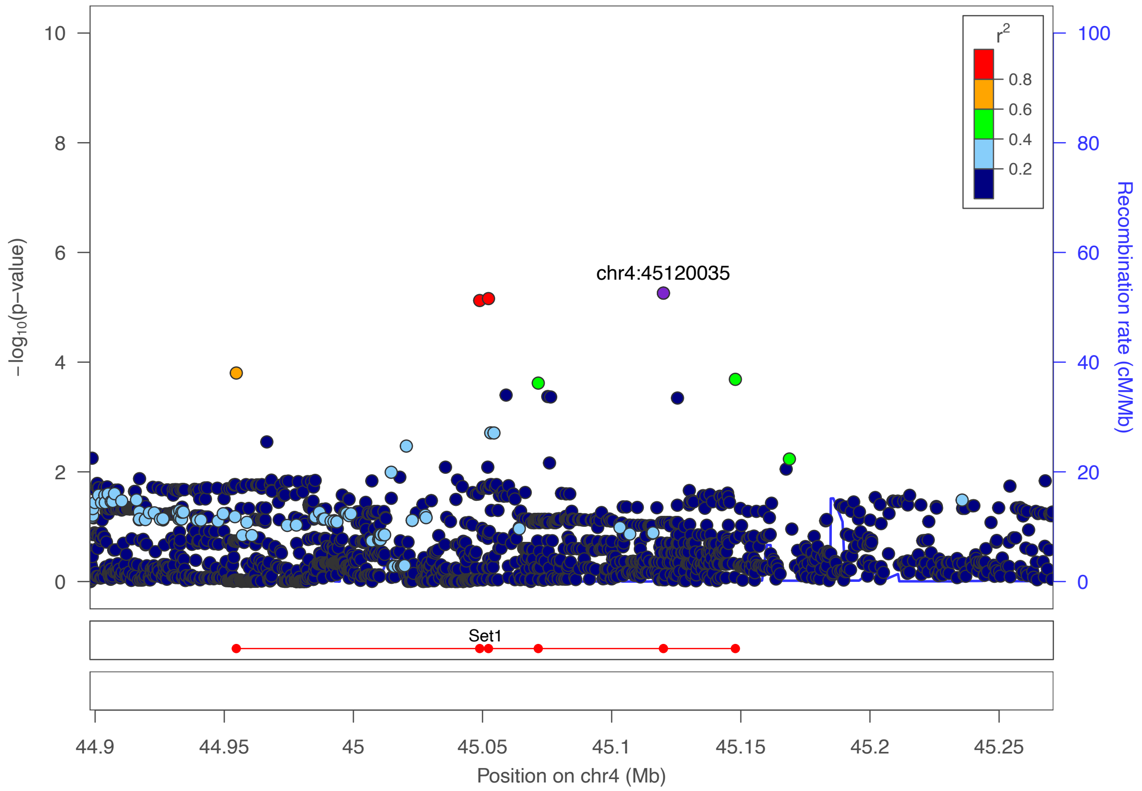

***Supplementary Figure 8:*** *Regional association plot for the locus 4p12 with the top hit chr4:45120035 (rs113429955). The purple dot represents the most strongly associated SNP with ulcerative colitis. The color of the dots represents the linkage disequilibrium (LD) with the most strongly associated SNP (see color legend). The positions represent the genome build GRCh38. The recombination rate is shown in centimorgans (cM) per million base pairs (Mb). The bottom part shows the name and locations of the genes within the region. The thicker blue line represents the position of the exons, while the thinner line represents the intronic regions. The direction of transcription is represented by an arrow behind the name of the gene. The plot was created using LocusZoom[2].*

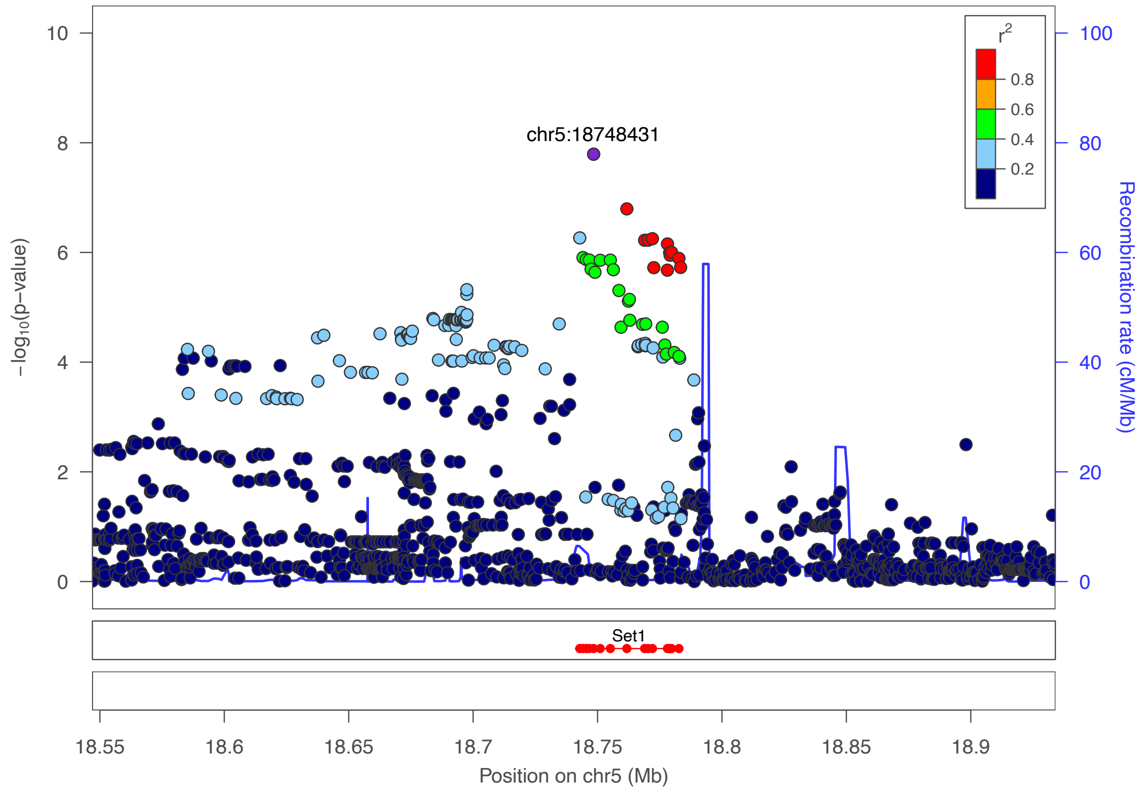

***Supplementary Figure 9:*** *Regional association plot for the locus 5p14.3 with the top hit chr5:18748431 (rs2937516). The purple dot represents the most strongly associated SNP with ulcerative colitis. The color of the dots represents the linkage disequilibrium (LD) with the most strongly associated SNP (see color legend). The positions represent the genome build GRCh38. The recombination rate is shown in centimorgans (cM) per million base pairs (Mb). The bottom part shows the name and locations of the genes within the region. The thicker blue line represents the position of the exons, while the thinner line represents the intronic regions. The direction of transcription is represented by an arrow behind the name of the gene. The plot was created using LocusZoom[2].*

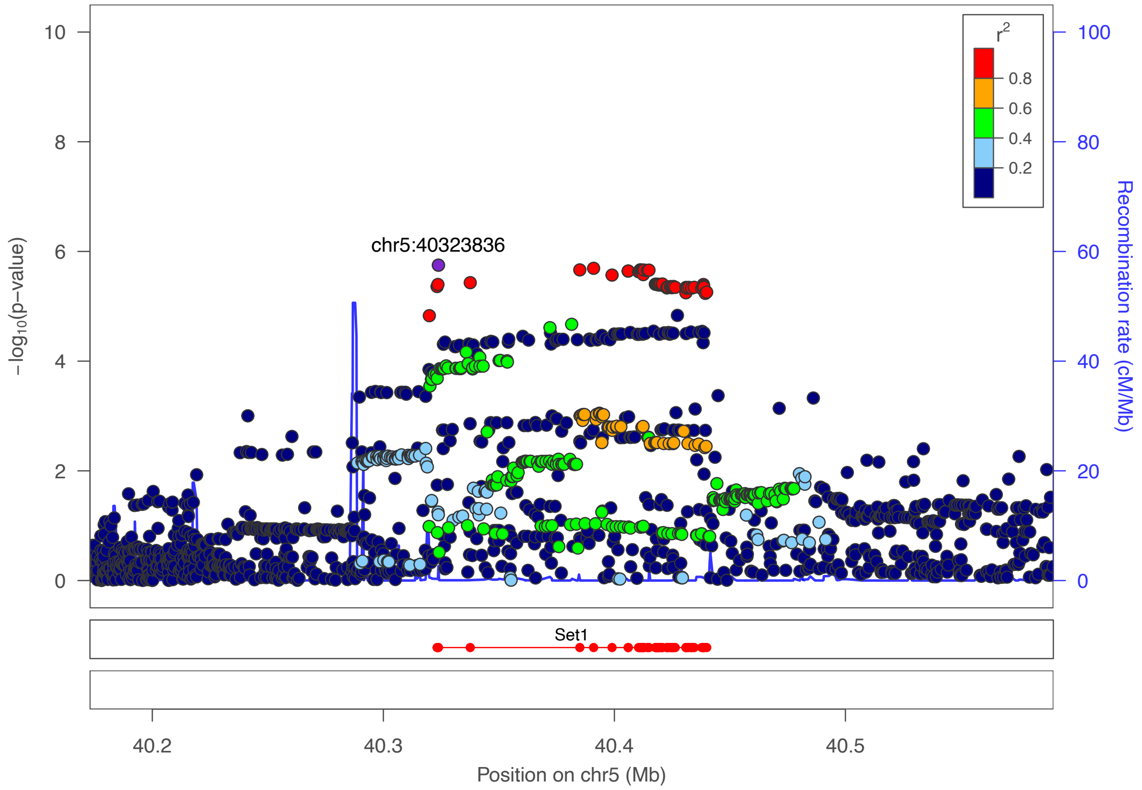

***Supplementary Figure 10:*** *Regional association plot for the locus 5p13.1 with the top hit chr5:40323836 (rs348594). The purple dot represents the most strongly associated SNP with ulcerative colitis. The color of the dots represents the linkage disequilibrium (LD) with the most strongly associated SNP (see color legend). The positions represent the genome build GRCh38. The recombination rate is shown in centimorgans (cM) per million base pairs (Mb). The bottom part shows the name and locations of the genes within the region. The thicker blue line represents the position of the exons, while the thinner line represents the intronic regions. The direction of transcription is represented by an arrow behind the name of the gene. The plot was created using LocusZoom[2].*

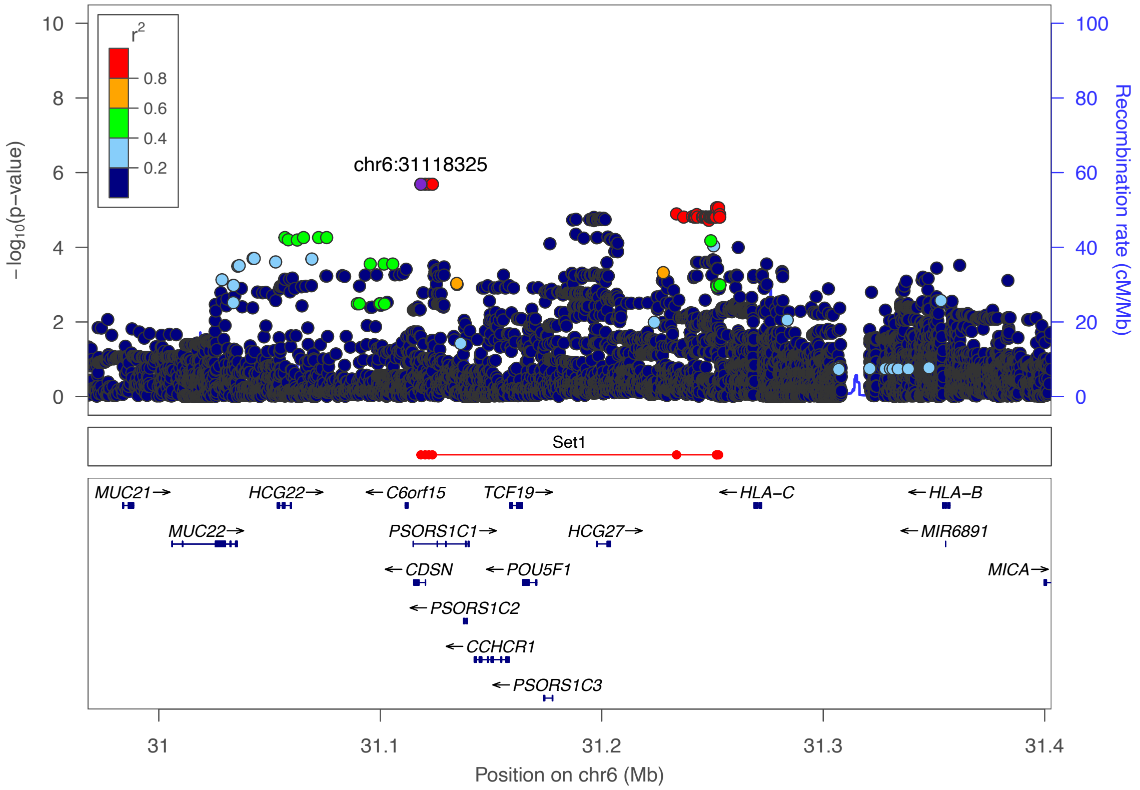

***Supplementary Figure 11:*** *Regional association plot for the locus 6p21.33 with the top hit chr6:31118325 (rs117198148). The purple dot represents the most strongly associated SNP with ulcerative colitis. The color of the dots represents the linkage disequilibrium (LD) with the most strongly associated SNP (see color legend). The positions represent the genome build GRCh38. The recombination rate is shown in centimorgans (cM) per million base pairs (Mb). The bottom part shows the name and locations of the genes within the region. The thicker blue line represents the position of the exons, while the thinner line represents the intronic regions. The direction of transcription is represented by an arrow behind the name of the gene. The plot was created using LocusZoom[2].*

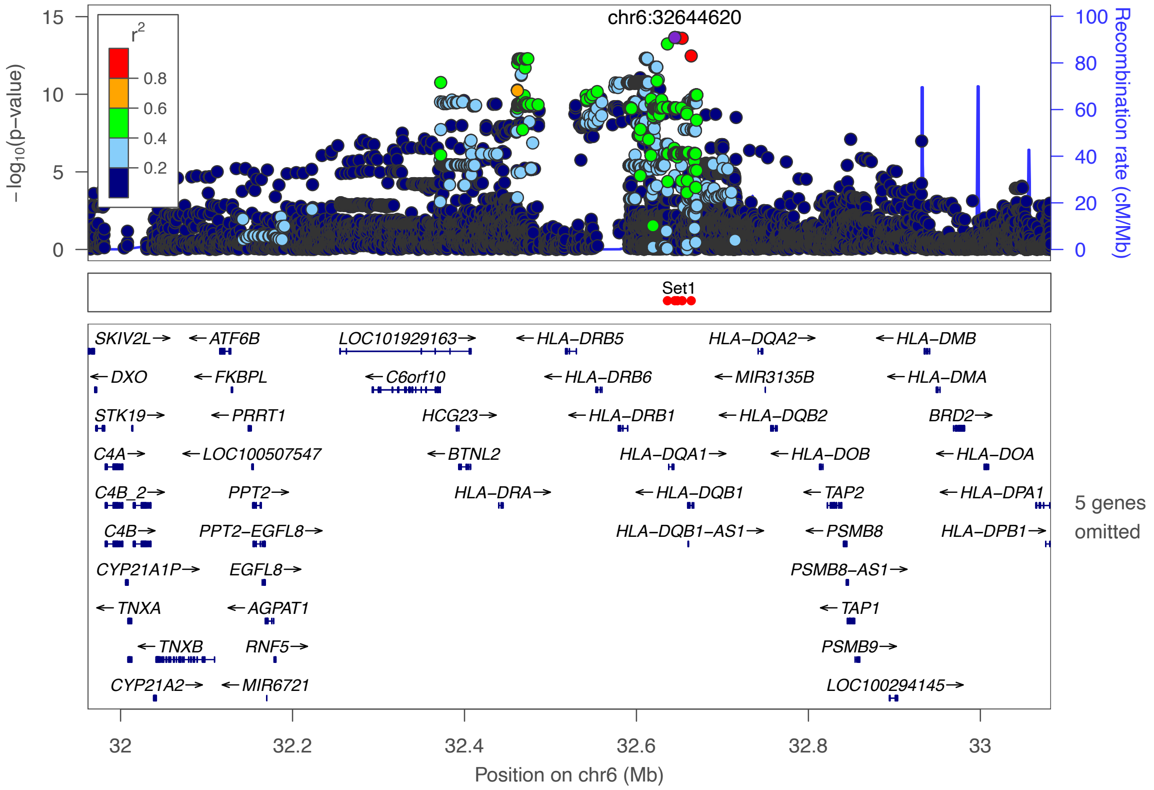

***Supplementary Figure 12:*** *Regional association plot for the locus 6p21.32 with the top hit chr6:32644620 (rs6927022). The purple dot represents the most strongly associated SNP with ulcerative colitis. The color of the dots represents the linkage disequilibrium (LD) with the most strongly associated SNP (see color legend). The positions represent the genome build GRCh38. The recombination rate is shown in centimorgans (cM) per million base pairs (Mb). The bottom part shows the name and locations of the genes within the region. The thicker blue line represents the position of the exons, while the thinner line represents the intronic regions. The direction of transcription is represented by an arrow behind the name of the gene. The plot was created using LocusZoom*[2].

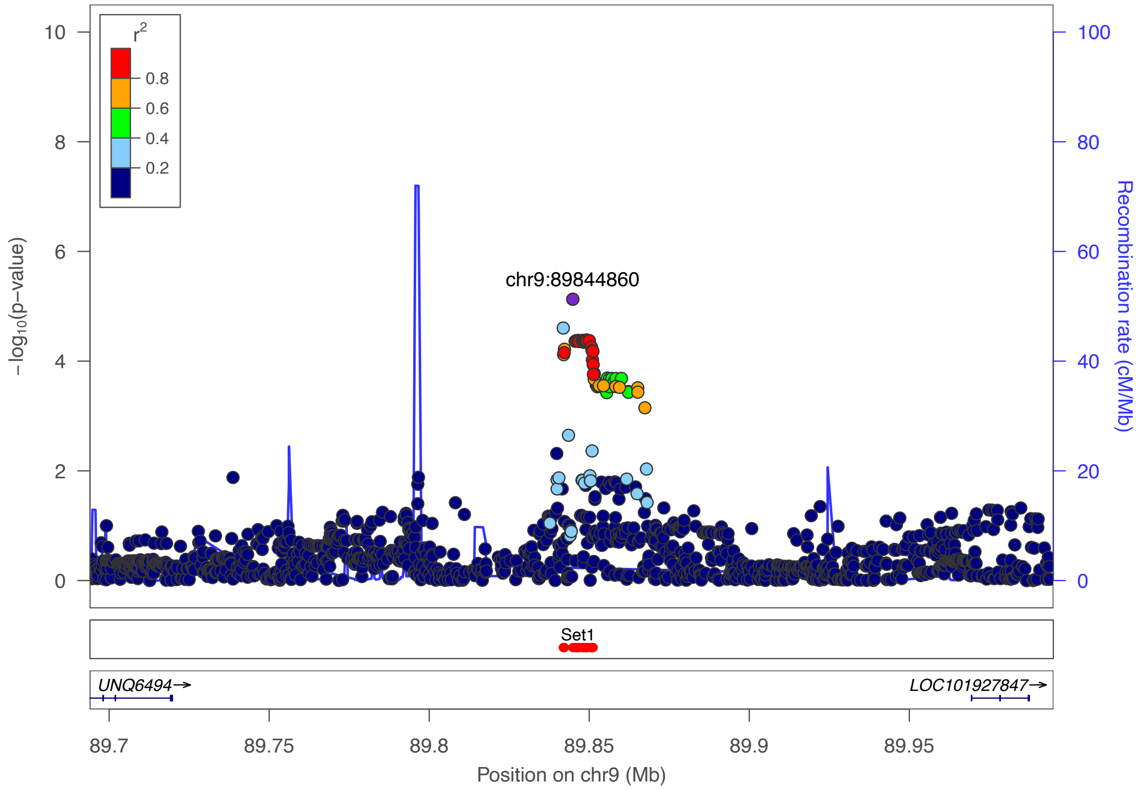

***Supplementary Figure 13:*** *Regional association plot for the locus 9q22.2 with the top hit chr9:89844860 (rs36147380). The purple dot represents the most strongly associated SNP with ulcerative colitis. The color of the dots represents the linkage disequilibrium (LD) with the most strongly associated SNP (see color legend). The positions represent the genome build GRCh38. The recombination rate is shown in centimorgans (cM) per million base pairs (Mb). The bottom part shows the name and locations of the genes within the region. The thicker blue line represents the position of the exons, while the thinner line represents the intronic regions. The direction of transcription is represented by an arrow behind the name of the gene. The plot was created using LocusZoom*[2]*.*

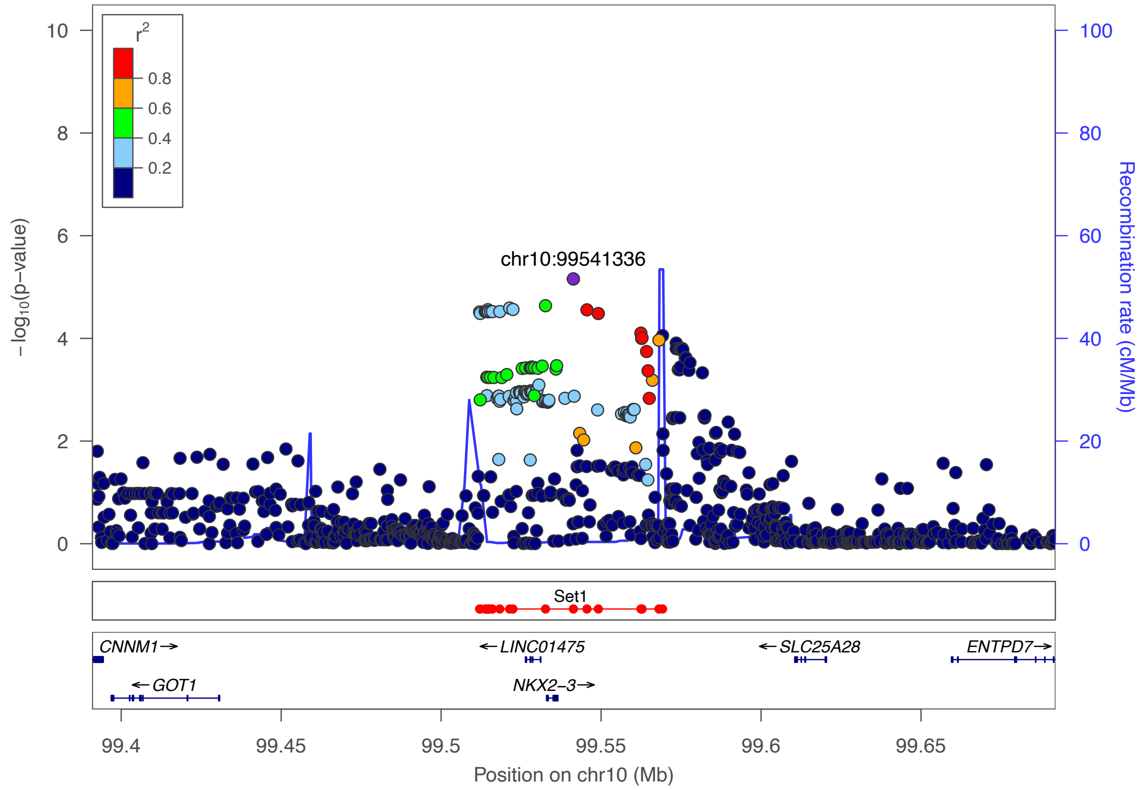

***Supplementary Figure 14:*** *Regional association plot for the locus 10q24.2 with the top hit chr10:99541336 (rs4590800). The purple dot represents the most strongly associated SNP with ulcerative colitis. The color of the dots represents the linkage disequilibrium (LD) with the most strongly associated SNP (see color legend). The positions represent the genome build GRCh38. The recombination rate is shown in centimorgans (cM) per million base pairs (Mb). The bottom part shows the name and locations of the genes within the region. The thicker blue line represents the position of the exons, while the thinner line represents the intronic regions. The direction of transcription is represented by an arrow behind the name of the gene. The plot was created using LocusZoom[2].*

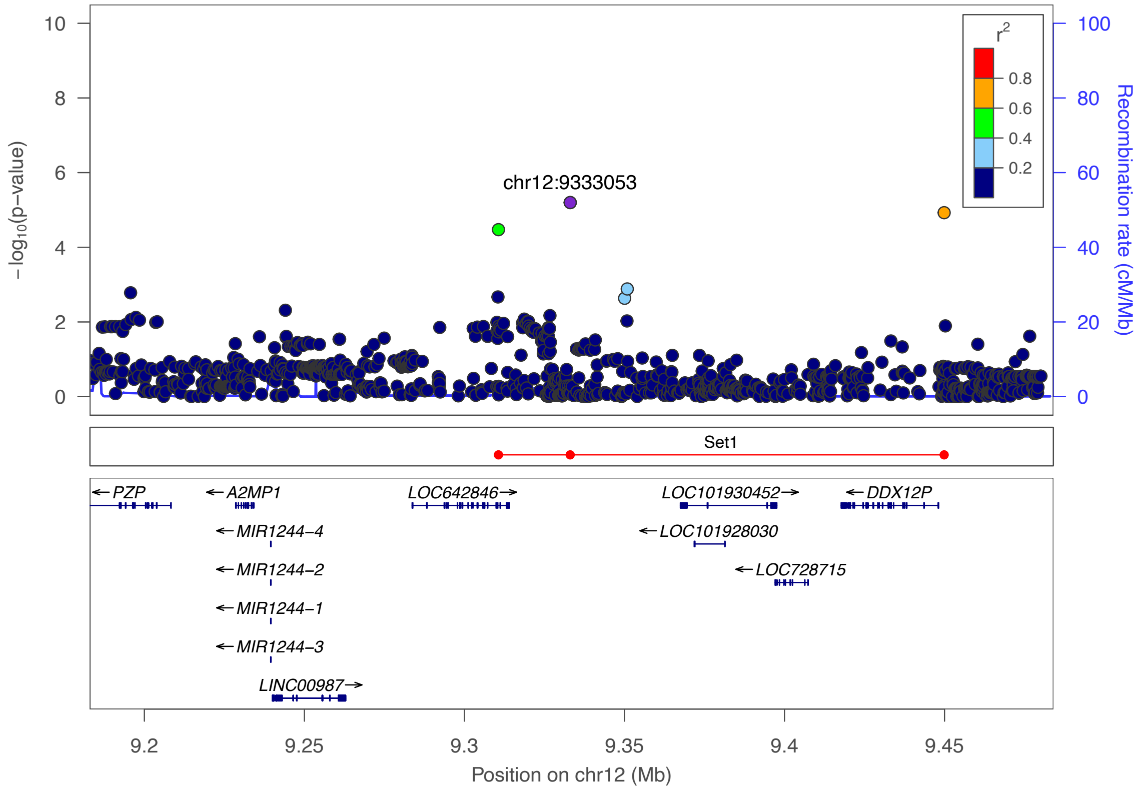

***Supplementary Figure 15:*** *Regional association plot for the locus 12p13.31 with the top hit chr12:9333053 (rs187033004). The purple dot represents the most strongly associated SNP with ulcerative colitis. The color of the dots represents the linkage disequilibrium (LD) with the most strongly associated SNP (see color legend). The positions represent the genome build GRCh38. The recombination rate is shown in centimorgans (cM) per million base pairs (Mb). The bottom part shows the name and locations of the genes within the region. The thicker blue line represents the position of the exons, while the thinner line represents the intronic regions. The direction of transcription is represented by an arrow behind the name of the gene. The plot was created using LocusZoom[2].*

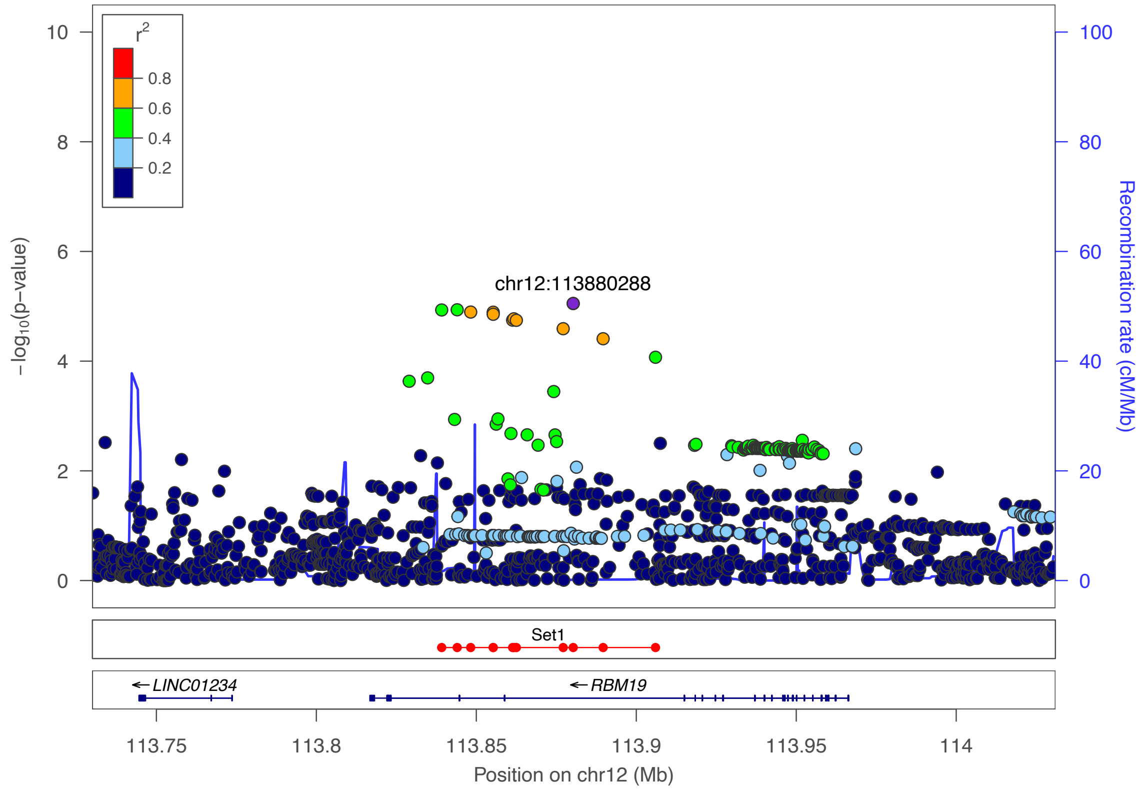

***Supplementary Figure 16:*** *Regional association plot for the locus 12q24.13 with the top hit chr12:113880288 (rs3782449). The purple dot represents the most strongly associated SNP with ulcerative colitis. The color of the dots represents the linkage disequilibrium (LD) with the most strongly associated SNP (see color legend). The positions represent the genome build GRCh38. The recombination rate is shown in centimorgans (cM) per million base pairs (Mb). The bottom part shows the name and locations of the genes within the region. The thicker blue line represents the position of the exons, while the thinner line represents the intronic regions. The direction of transcription is represented by an arrow behind the name of the gene. The plot was created using LocusZoom[2].*

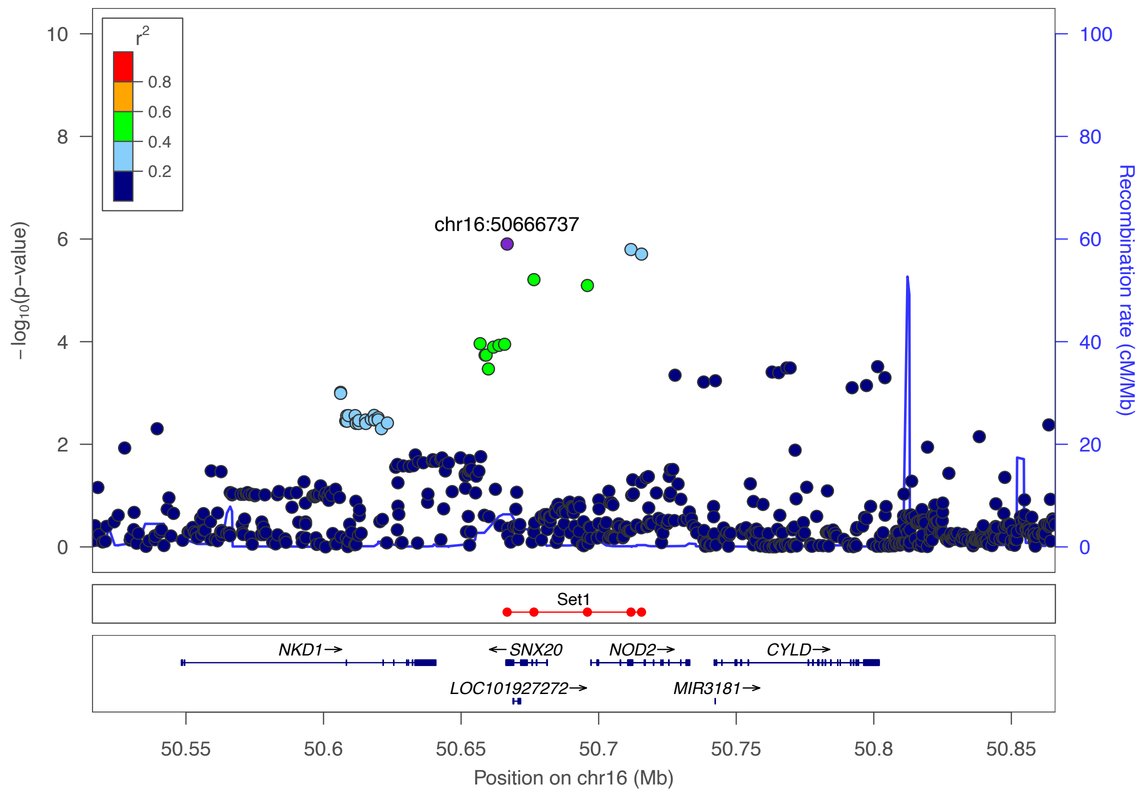

***Supplementary Figure 17:*** *Regional association plot for the locus 16q12.1 with the top hit chr16:50666737 (rs139397276). The purple dot represents the most strongly associated SNP with ulcerative colitis. The color of the dots represents the linkage disequilibrium (LD) with the most strongly associated SNP (see color legend). The positions represent the genome build GRCh38. The recombination rate is shown in centimorgans (cM) per million base pairs (Mb). The bottom part shows the name and locations of the genes within the region. The thicker blue line represents the position of the exons, while the thinner line represents the intronic regions. The direction of transcription is represented by an arrow behind the name of the gene. The plot was created using LocusZoom*[2]*.*

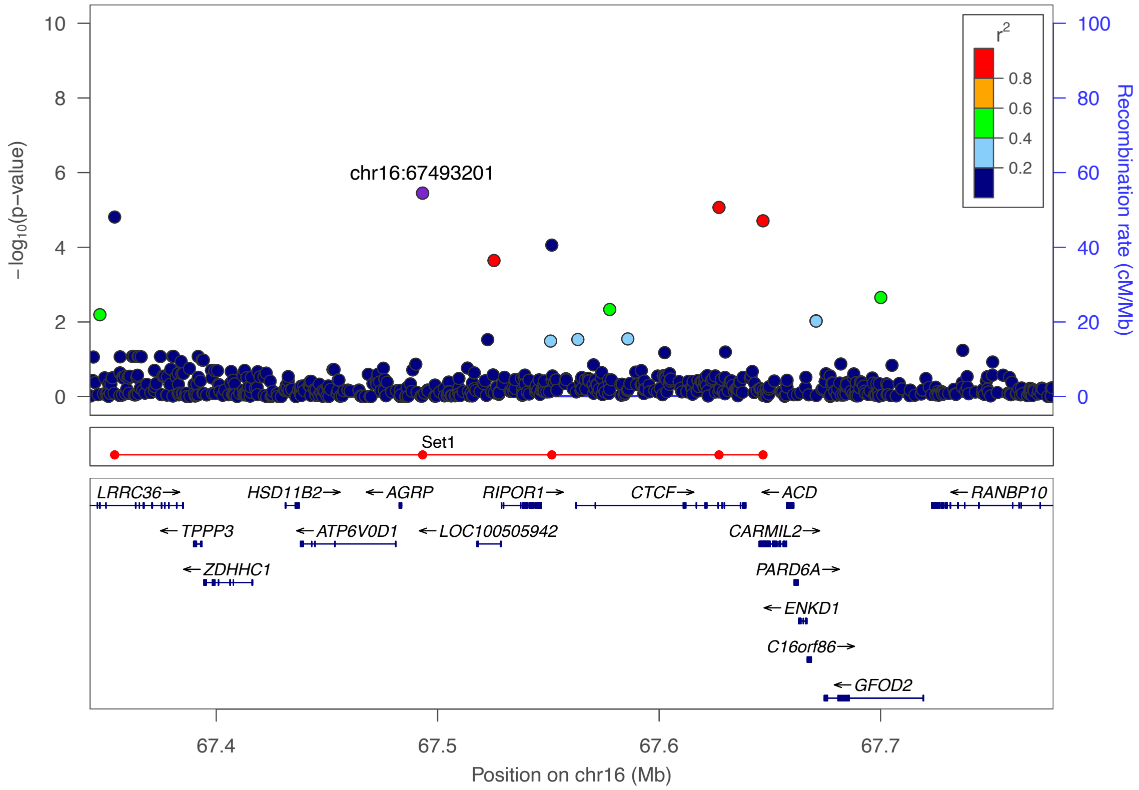

***Supplementary Figure 18:*** *Regional association plot for the locus 16q22.1 with the top hit chr16:67493201 (rs77919558). The purple dot represents the most strongly associated SNP with ulcerative colitis. The color of the dots represents the linkage disequilibrium (LD) with the most strongly associated SNP (see color legend). The positions represent the genome build GRCh38. The recombination rate is shown in centimorgans (cM) per million base pairs (Mb). The bottom part shows the name and locations of the genes within the region. The thicker blue line represents the position of the exons, while the thinner line represents the intronic regions. The direction of transcription is represented by an arrow behind the name of the gene. The plot was created using LocusZoom[2].*

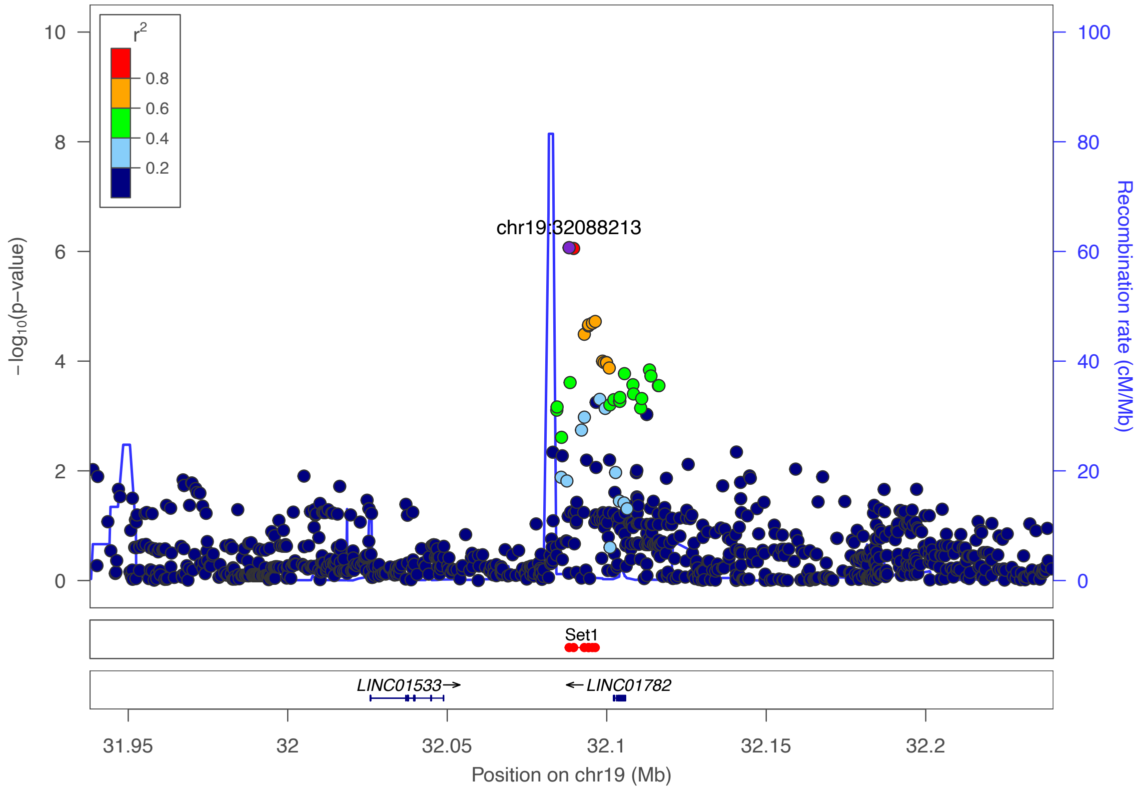

***Supplementary Figure 19:*** *Regional association plot for the locus 19q13.11 with the top hit chr19:32088213 (rs6510221). The purple dot represents the most strongly associated SNP with ulcerative colitis. The color of the dots represents the linkage disequilibrium (LD) with the most strongly associated SNP (see color legend). The positions represent the genome build GRCh38. The recombination rate is shown in centimorgans (cM) per million base pairs (Mb). The bottom part shows the name and locations of the genes within the region. The thicker blue line represents the position of the exons, while the thinner line represents the intronic regions. The direction of transcription is represented by an arrow behind the name of the gene. The plot was created using LocusZoom[2].*

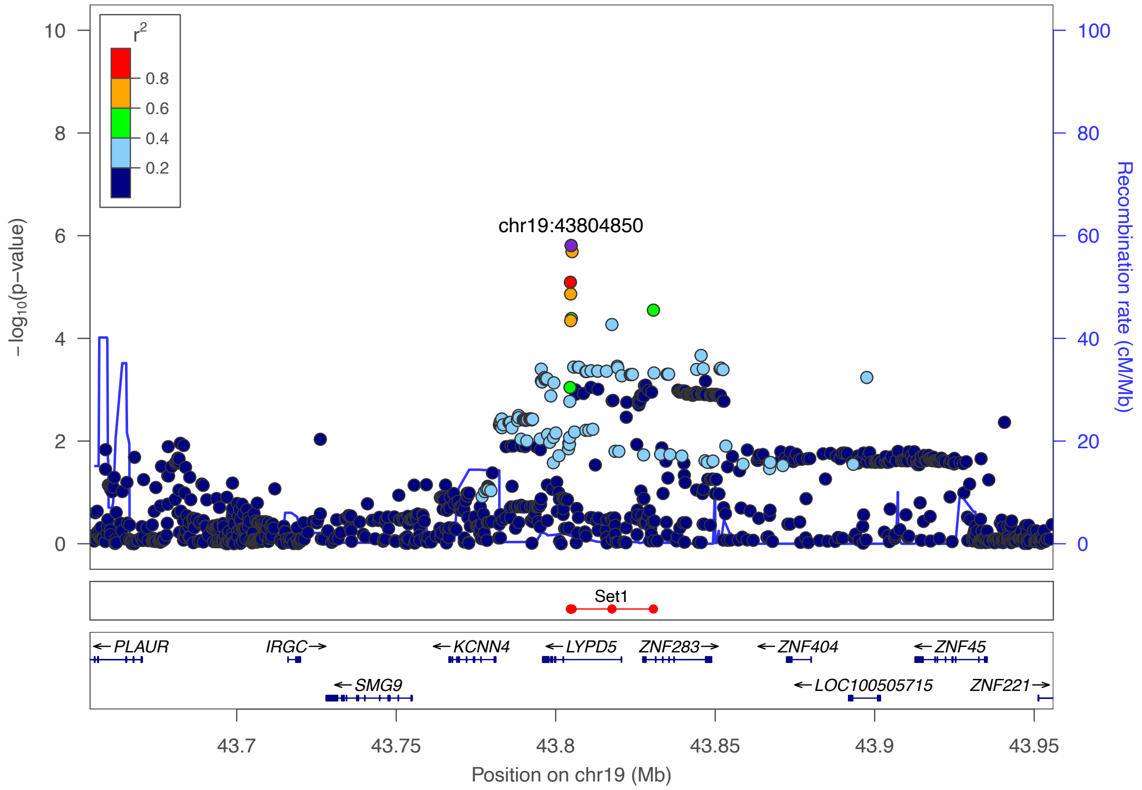

***Supplementary Figure 20:*** *Regional association plot for the locus 19q13.31 with the top hit chr19:43804850 (rs364691). The purple dot represents the most strongly associated SNP with ulcerative colitis. The color of the dots represents the linkage disequilibrium (LD) with the most strongly associated SNP (see color legend). The positions represent the genome build GRCh38. The recombination rate is shown in centimorgans (cM) per million base pairs (Mb). The bottom part shows the name and locations of the genes within the region. The thicker blue line represents the position of the exons, while the thinner line represents the intronic regions. The direction of transcription is represented by an arrow behind the name of the gene. The plot was created using LocusZoom*[2].

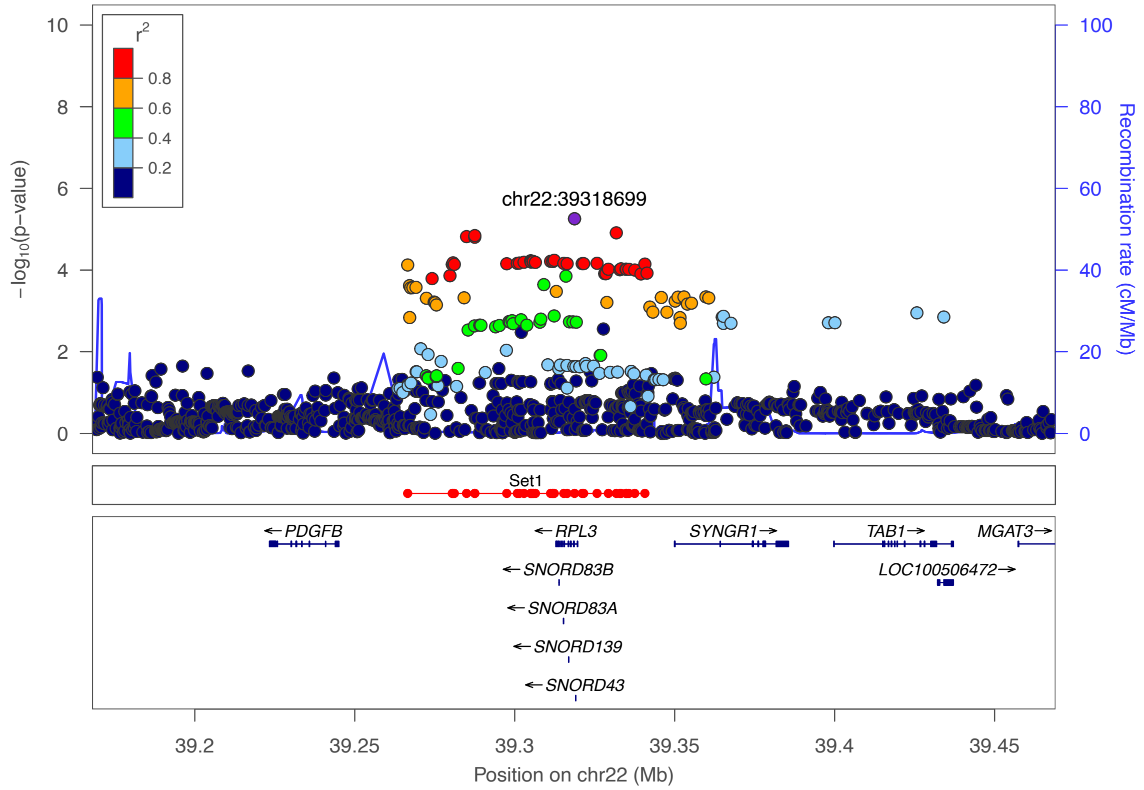

***Supplementary Figure 21:*** *Regional association plot for the locus 22q13.1 with the top hit chr22:39318699 (rs1569498). The purple dot represents the most strongly associated SNP with ulcerative colitis. The color of the dots represents the linkage disequilibrium (LD) with the most strongly associated SNP (see color legend). The positions represent the genome build GRCh38. The recombination rate is shown in centimorgans (cM) per million base pairs (Mb). The bottom part shows the name and locations of the genes within the region. The thicker blue line represents the position of the exons, while the thinner line represents the intronic regions. The direction of transcription is represented by an arrow behind the name of the gene. The plot was created using LocusZoom[2].*

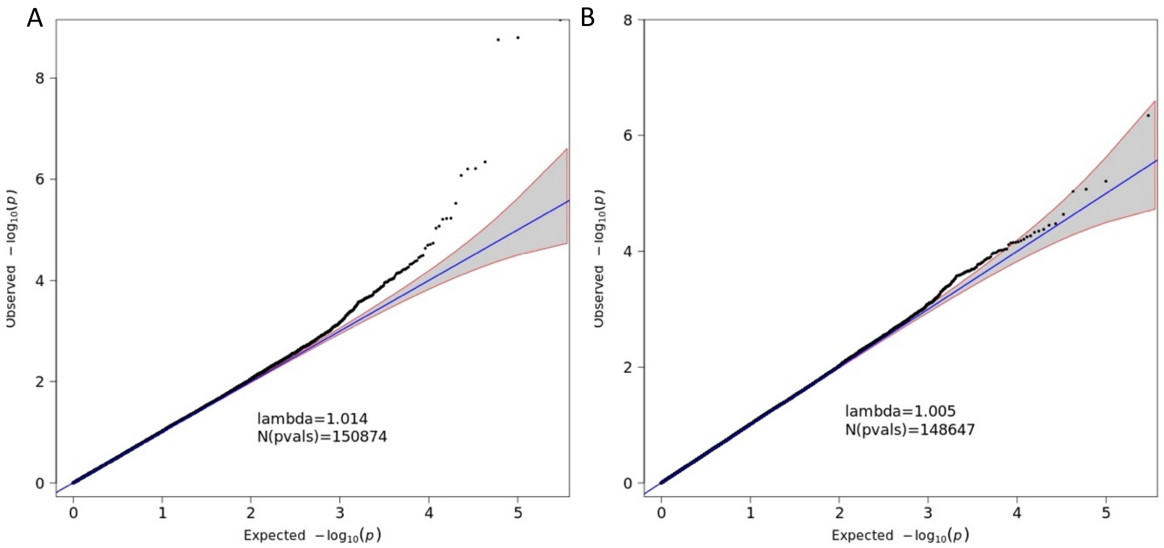

***Supplementary Figure 22:*** *Quantile-quantile plot of association summary statistics of the whole exome data. The 95% concentration band under random sampling is shown in gray. The genomic inflation factor lambda is defined as the ratio of the medians of the sample $\chi 2$ test statistics and the 1-df $\chi 2$ distribution (0.455).[1] The left figure includes all 150,874 variants with MAF > 1 % and an imputation score $r^2$>0.6. The right figure excludes the variants of the HLA-region (chr6:29-34MB).*

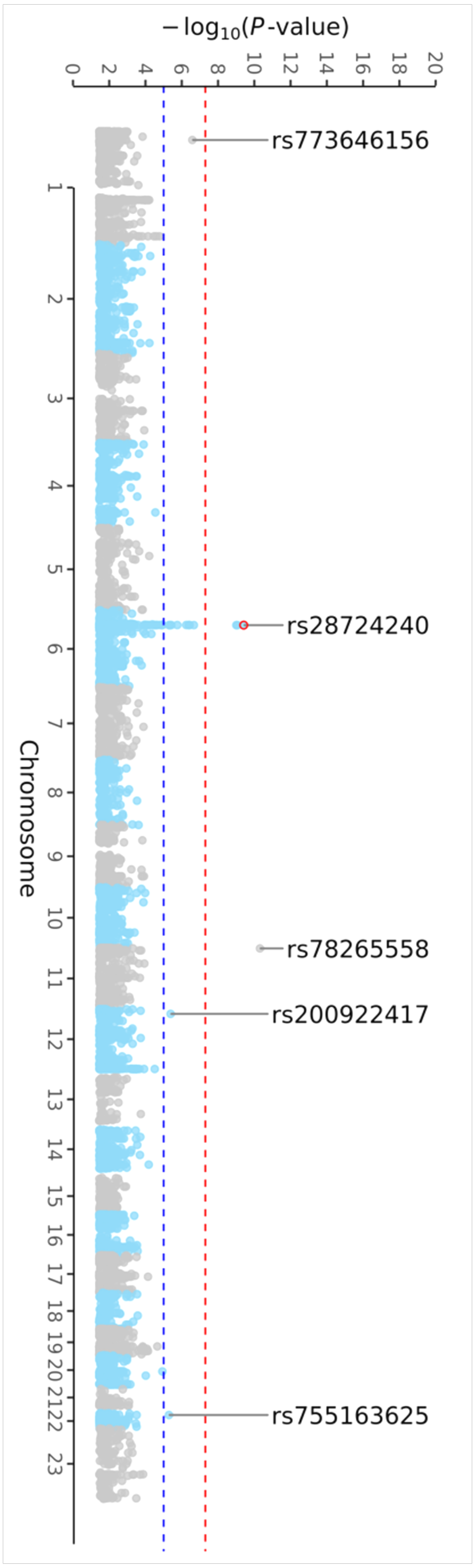

***Supplementary Figure 23:*** *Manhattan plot of the exome data with a MAF >1% and an imputation score r2 >0.6. All loci of at least nominal significance (blue horizontal line; P<1×10-5) are annotated by the SNP-ID. Loci with LD support are highlighted with a blue (nominal significance) or red circle (genome-wide significance, red horizontal line; P<5×10$^{8}$).*

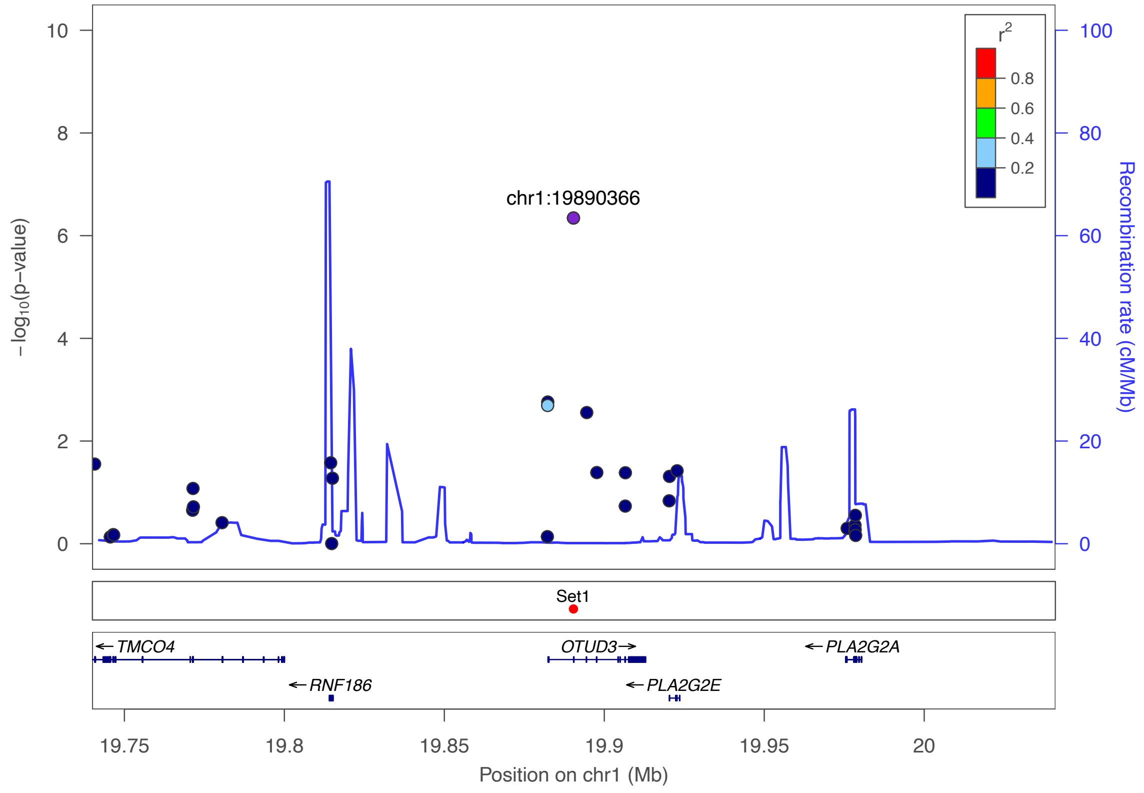

***Supplementary Figure 24:*** *Regional association plot for the locus 1p36.13 in the exome data with the top hit chr1:19890366 (rs7523442). The purple dot represents the most strongly associated SNP with ulcerative colitis. The color of the dots represents the linkage disequilibrium (LD) with the most strongly associated SNP (see color legend). The positions represent the genome build GRCh38. The recombination rate is shown in centimorgans (cM) per million base pairs (Mb). The bottom part shows the name and locations of the genes within the region. The thicker blue line represents the position of the exons, while the thinner line represents the intronic regions. The direction of transcription is represented by an arrow behind the name of the gene. The plot was created using LocusZoom[2].*

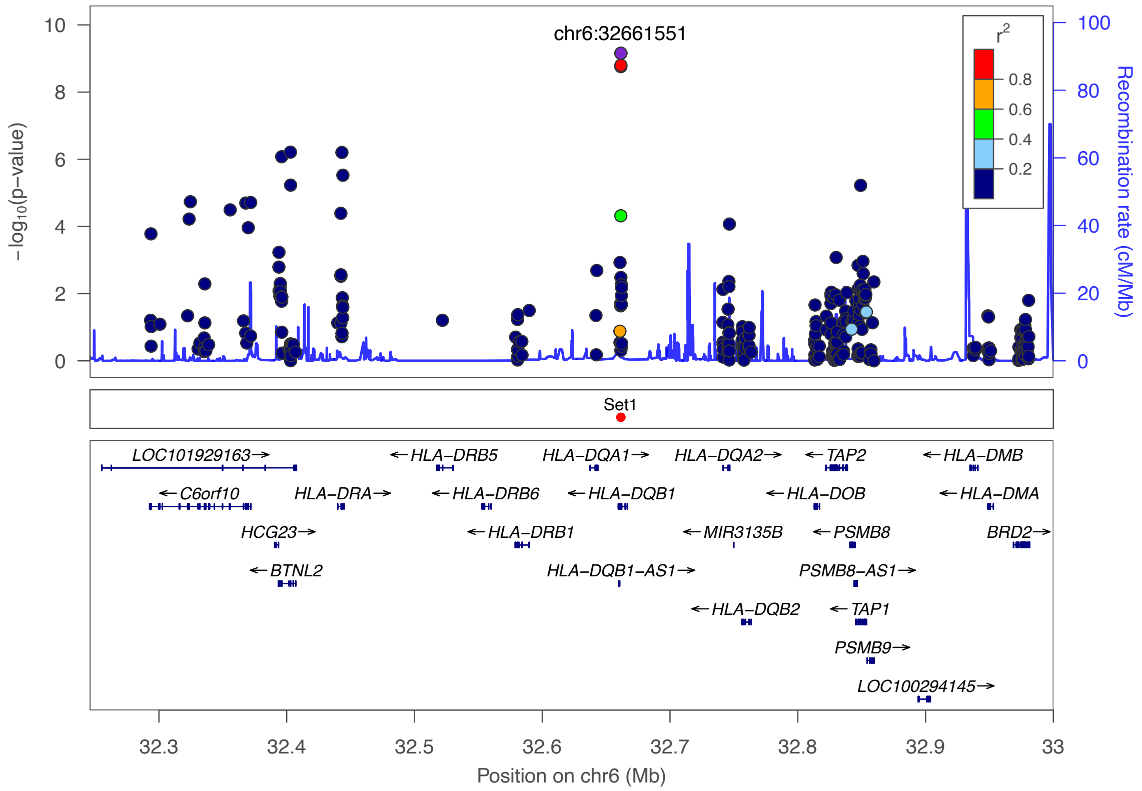

***Supplementary Figure 25:*** *Regional association plot for the locus 6p21.32 in the exome data with the top hit chr6:32661551 (rs28724240). The purple dot represents the most strongly associated SNP with ulcerative colitis. The color of the dots represents the linkage disequilibrium (LD) with the most strongly associated SNP (see color legend). The positions represent the genome build GRCh38. The recombination rate is shown in centimorgans (cM) per million base pairs (Mb). The bottom part shows the name and locations of the genes within the region. The thicker blue line represents the position of the exons, while the thinner line represents the intronic regions. The direction of transcription is represented by an arrow behind the name of the gene. The plot was created using LocusZoom*[2]*.*

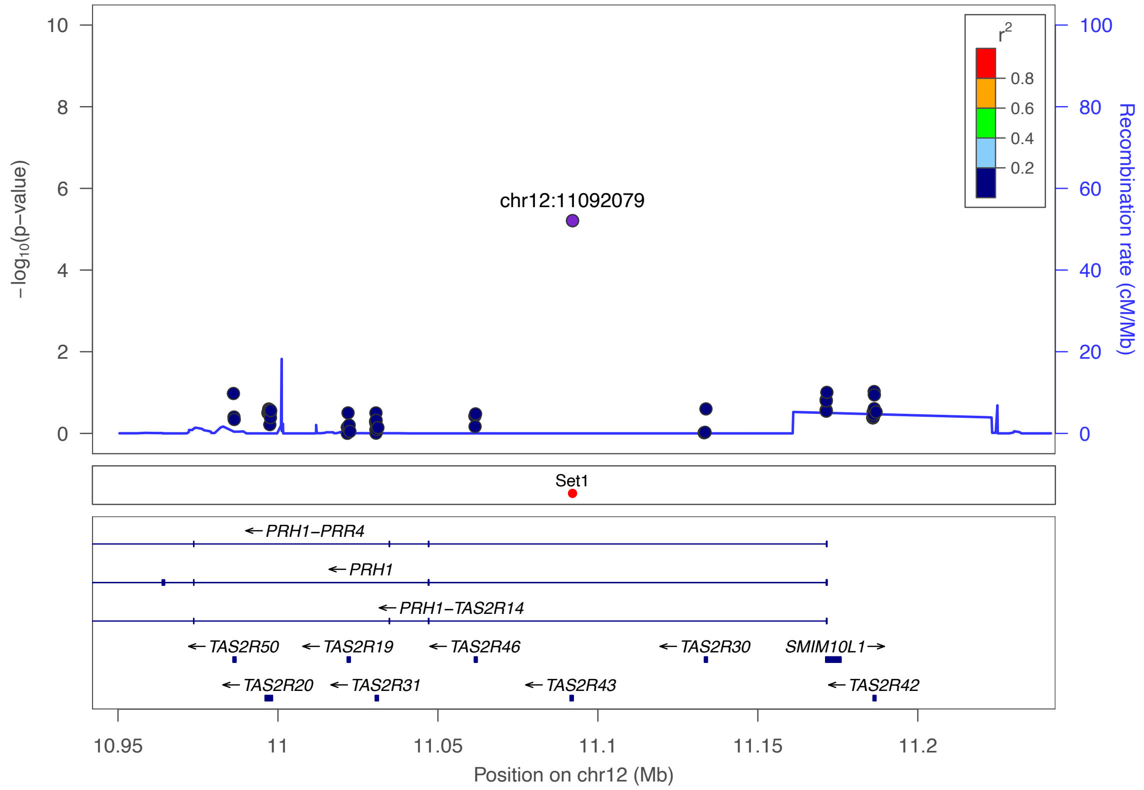

***Supplementary Figure 26:*** *Regional association plot for the locus 12p13.2 in the exome data with the top hit chr12:11092079 (rs113197337). The purple dot represents the most strongly associated SNP with ulcerative colitis. The color of the dots represents the linkage disequilibrium (LD) with the most strongly associated SNP (see color legend). The positions represent the genome build GRCh38. The recombination rate is shown in centimorgans (cM) per million base pairs (Mb). The bottom part shows the name and locations of the genes within the region. The thicker blue line represents the position of the exons, while the thinner line represents the intronic regions. The direction of transcription is represented by an arrow behind the name of the gene. The plot was created using LocusZoom[2].*

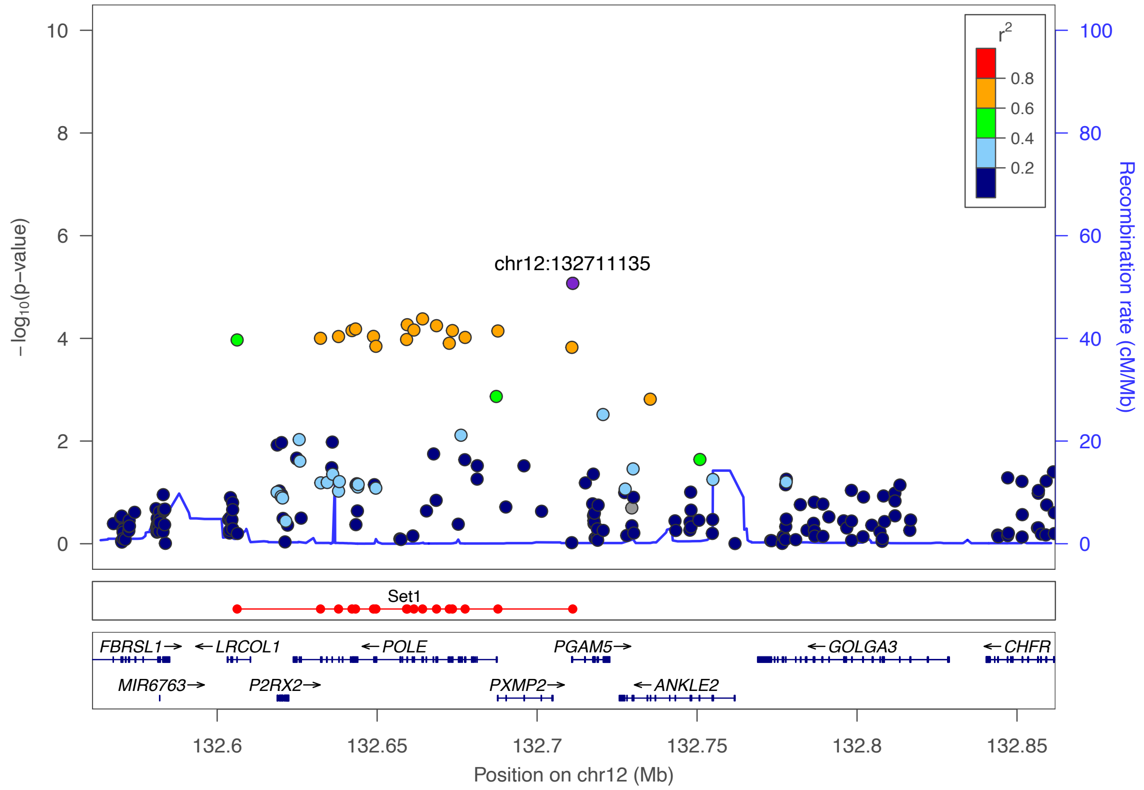

***Supplementary Figure 27:*** *Regional association plot for the locus 12q24.33 in the exome data with the top hit chr12:132711135 (rs7973452). The purple dot represents the most strongly associated SNP with ulcerative colitis. The color of the dots represents the linkage disequilibrium (LD) with the most strongly associated SNP (see color legend). The positions represent the genome build GRCh38. The recombination rate is shown in centimorgans (cM) per million base pairs (Mb). The bottom part shows the name and locations of the genes within the region. The thicker blue line represents the position of the exons, while the thinner line represents the intronic regions. The direction of transcription is represented by an arrow behind the name of the gene. The plot was created using LocusZoom[2].*

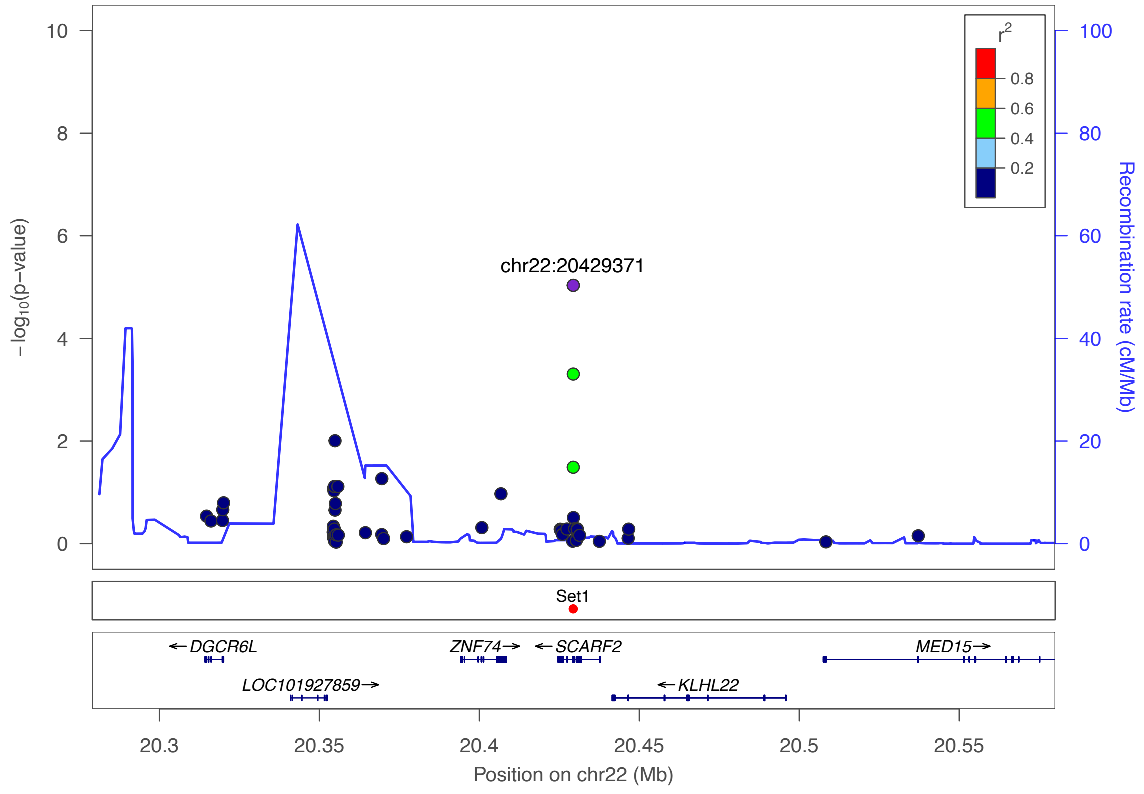

***Supplementary Figure 28:*** *Regional association plot for the locus 22q11.21 in the exome data with the top hit chr22:20429371 (rs755163625). The purple dot represents the most strongly associated SNP with ulcerative colitis. The color of the dots represents the linkage disequilibrium (LD) with the most strongly associated SNP (see color legend). The positions represent the genome build GRCh38. The recombination rate is shown in centimorgans (cM) per million base pairs (Mb). The bottom part shows the name and locations of the genes within the region. The thicker blue line represents the position of the exons, while the thinner line represents the intronic regions. The direction of transcription is represented by an arrow behind the name of the gene. The plot was created using LocusZoom[2].*

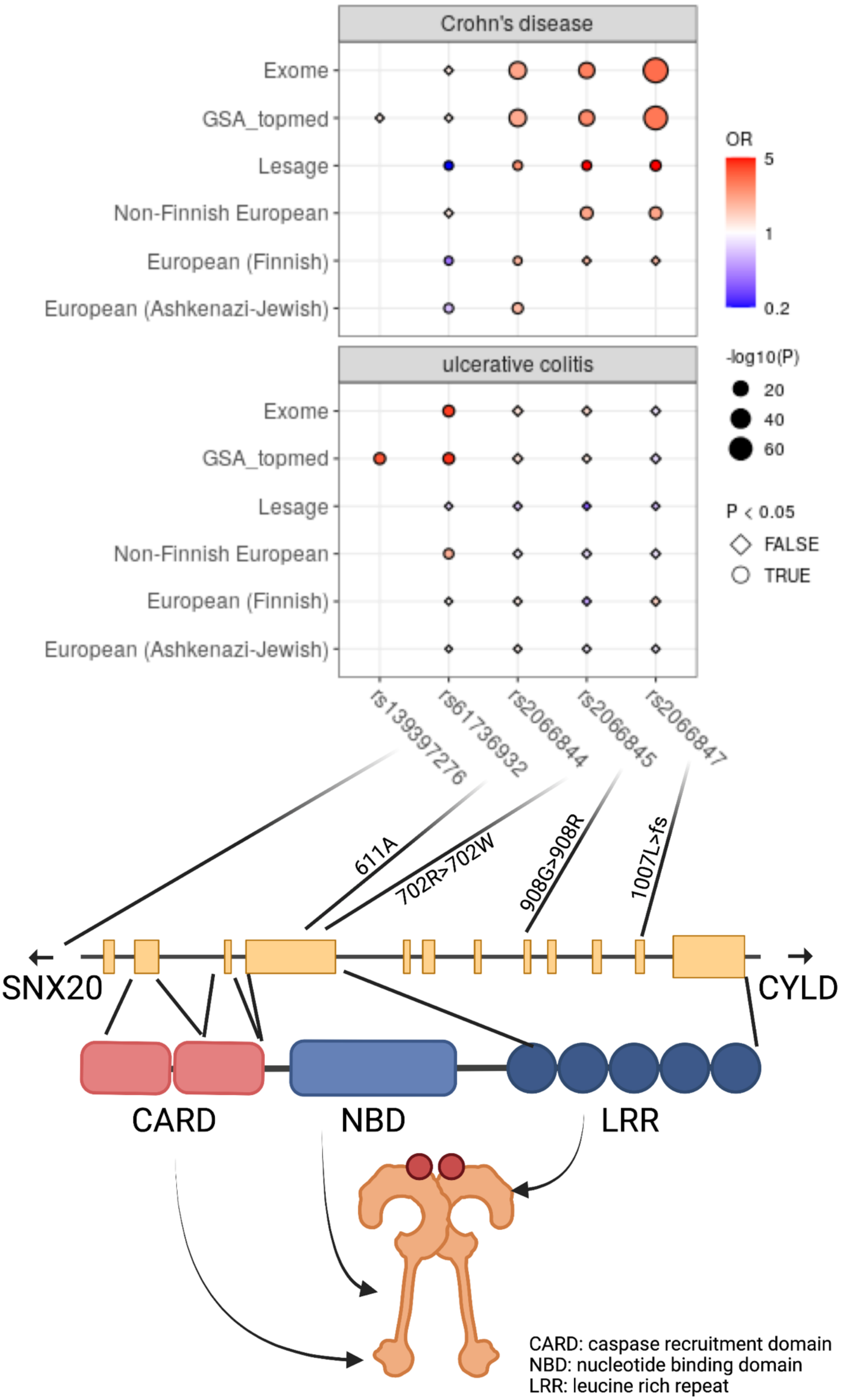
Crohn's disease
Exome
GSA_topmed
Lesage
Non-Finnish European
European (Finnish)
European (Ashkenazi-Jewish)
ulcerative colitis
OR
5
1
0.2
-log10(P)
20
40
60
P < 0.05
FALSE
TRUE
rs139397276
rs61736932
rs2066844
rs2066845
rs2066847
611A
702R>702W
908G>908R
1007L>fs
SNX20
CYLD
CARD
NBD
LRR
CARD: caspase recruitment domain
NBD: nucleotide binding domain
LRR: leucine rich repeat

***Supplementary Figure 29:*** *Associations at the NOD2 locus and the influence on the protein level. Rs139397277 is our main signal and rs61736932 the strongest associated variant within the exome data while the other three variants are those previously identified as associated with CD. Created with BiorRender.com.*

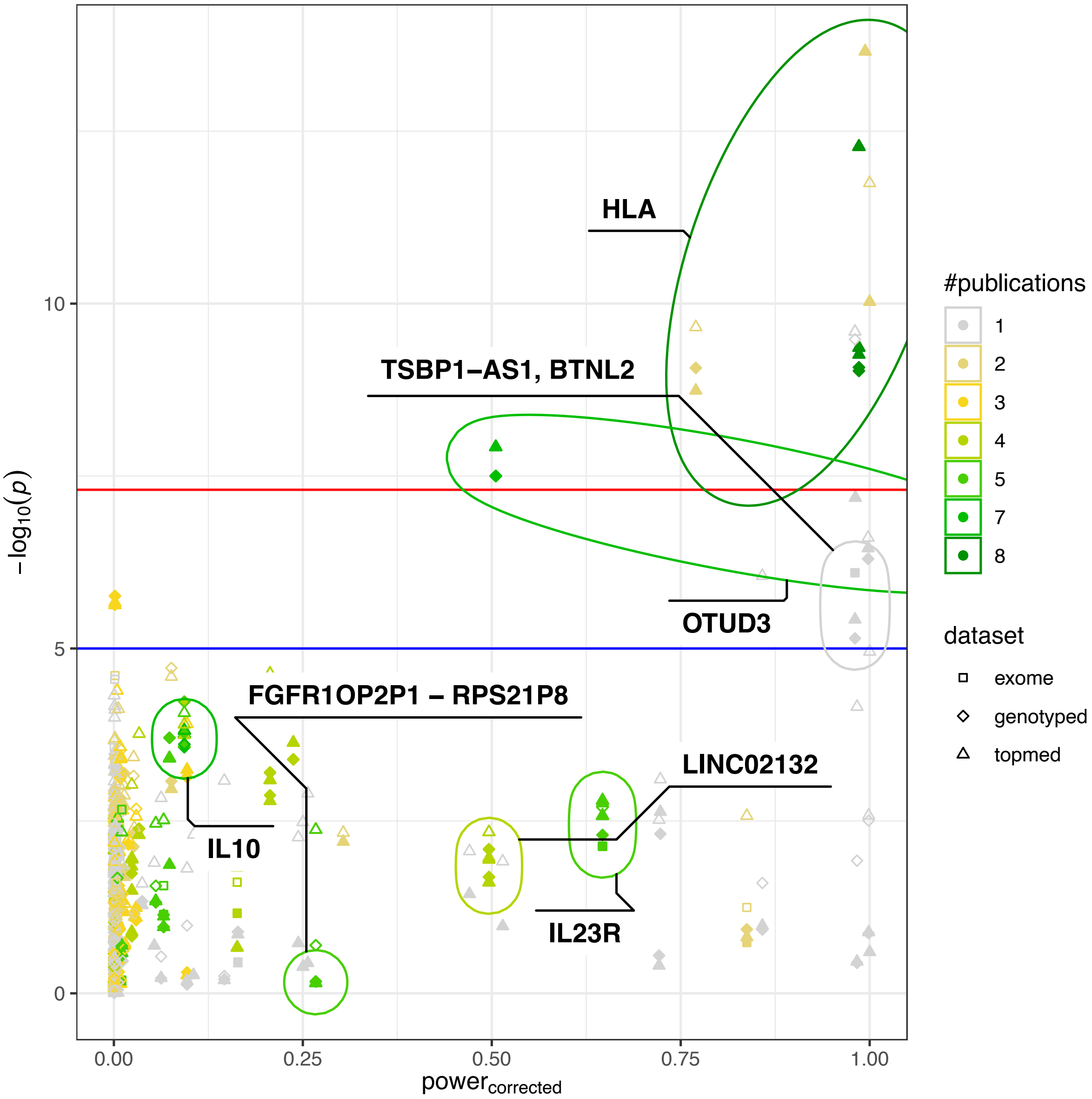

***Supplementary Figure 30:*** *Power analysis based on the GWAS catalog data. The corrected power is calculated based on the median odds ratios and standard errors as listed in the GWAS catalog, when removing the strongest effect if more than one association is given. The power is calculated for the nominal significance level $1 \times 10^{-5}$ and the frequencies given in our data.*

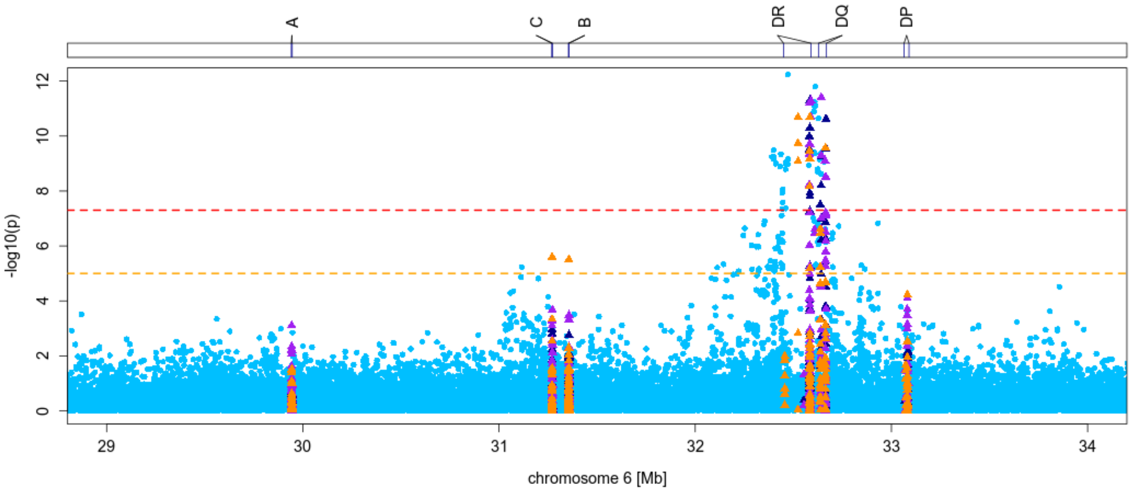

***Supplementary Figure 31:*** *Finemapping of the HLA region including the imputed alleles (orange), amino acids (purple) and nucleotides (dark blue) generated from the HIBAG imputation. The light blue dots in the background are the topmed imputed variants.*

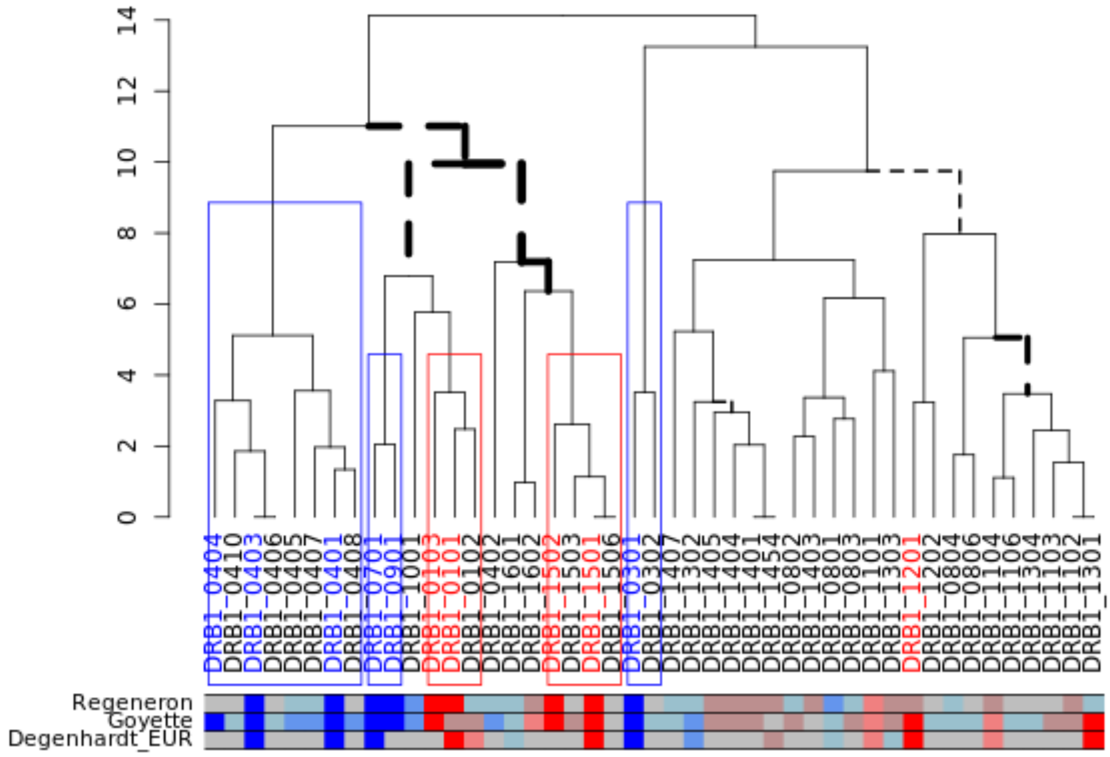

***Supplementary Figure 32:*** *Dendrogram of HLA-DRB1 alleles. The distances within the dendrogram are based on the Pearsons' correlation coefficient between the predicted eluted ligand mass spectrometry score from NetMHCIIpan-4.0. The main clusters of risk and protective alleles are highlighted. The colored box below shows the effect direction shown in our data as well as in the two most important publications on HLA in UC (blue: protective, red: risk, grey: no information, shading based on the p-value: grayish red/blue: no significance, mid red/blue: $p<0.05$, pure red/blue: $p<5 \times 10^{-8}$). All straight lines were also stable on other prediction subsets of the human proteome, while dotted lines represent lines that differ dependent on the proteome.*

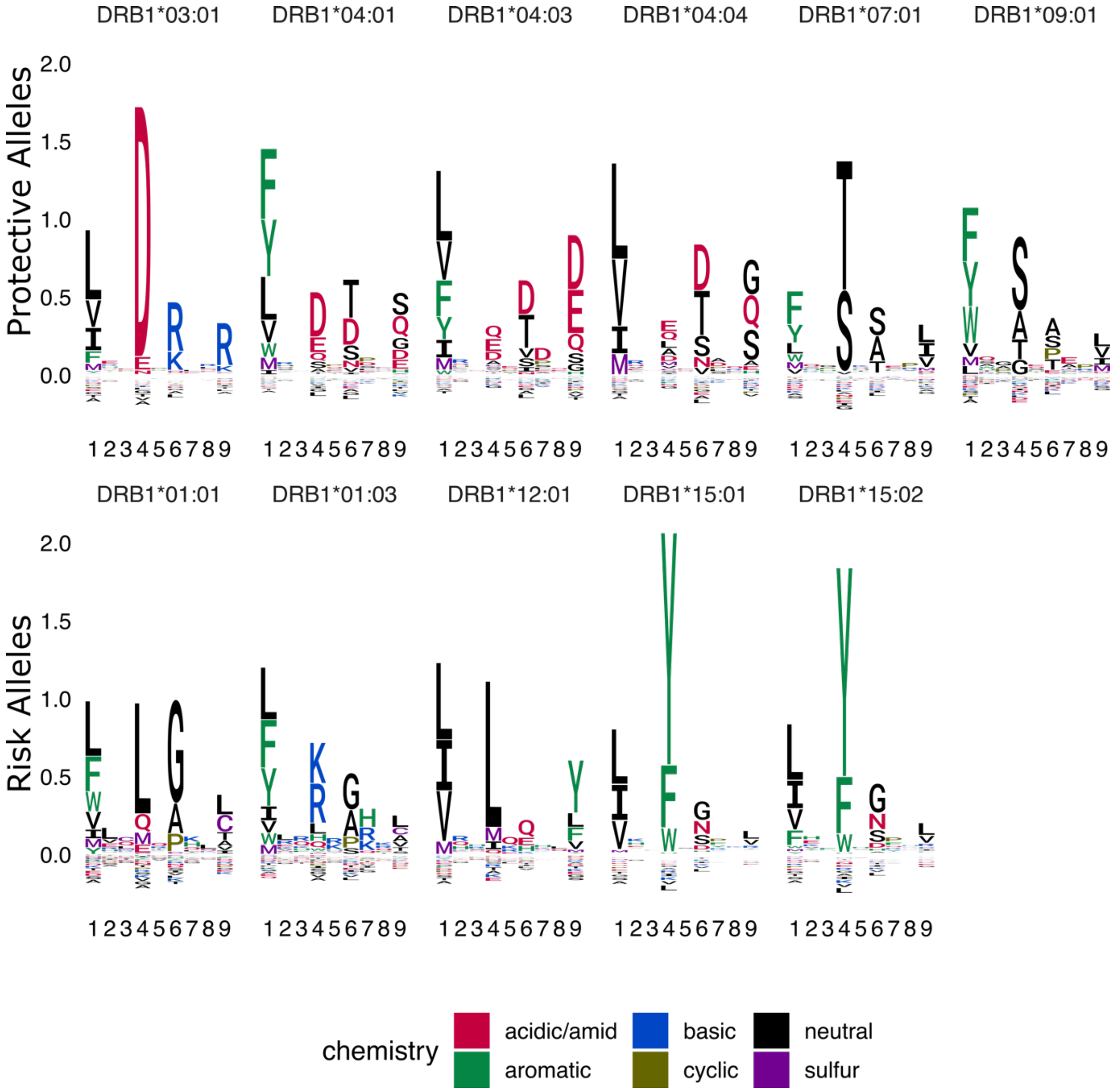

***Supplementary Figure 33:*** *Binding logo plot of associated HLA-DR alleles in differentiation to alleles with the other direction of effect. The upper row represents the protective associated alleles, the bottom line the logos of the risk alleles. The motifs are based on the NetMHCIIpan-4.0 predictions of the binding cores of all peptides at least annotated as weak binders, excluding peptides binding against one of the alleles of different direction of effect. The single letters represent the one letter amino acid code colored by the chemical properties of the amino acids.*

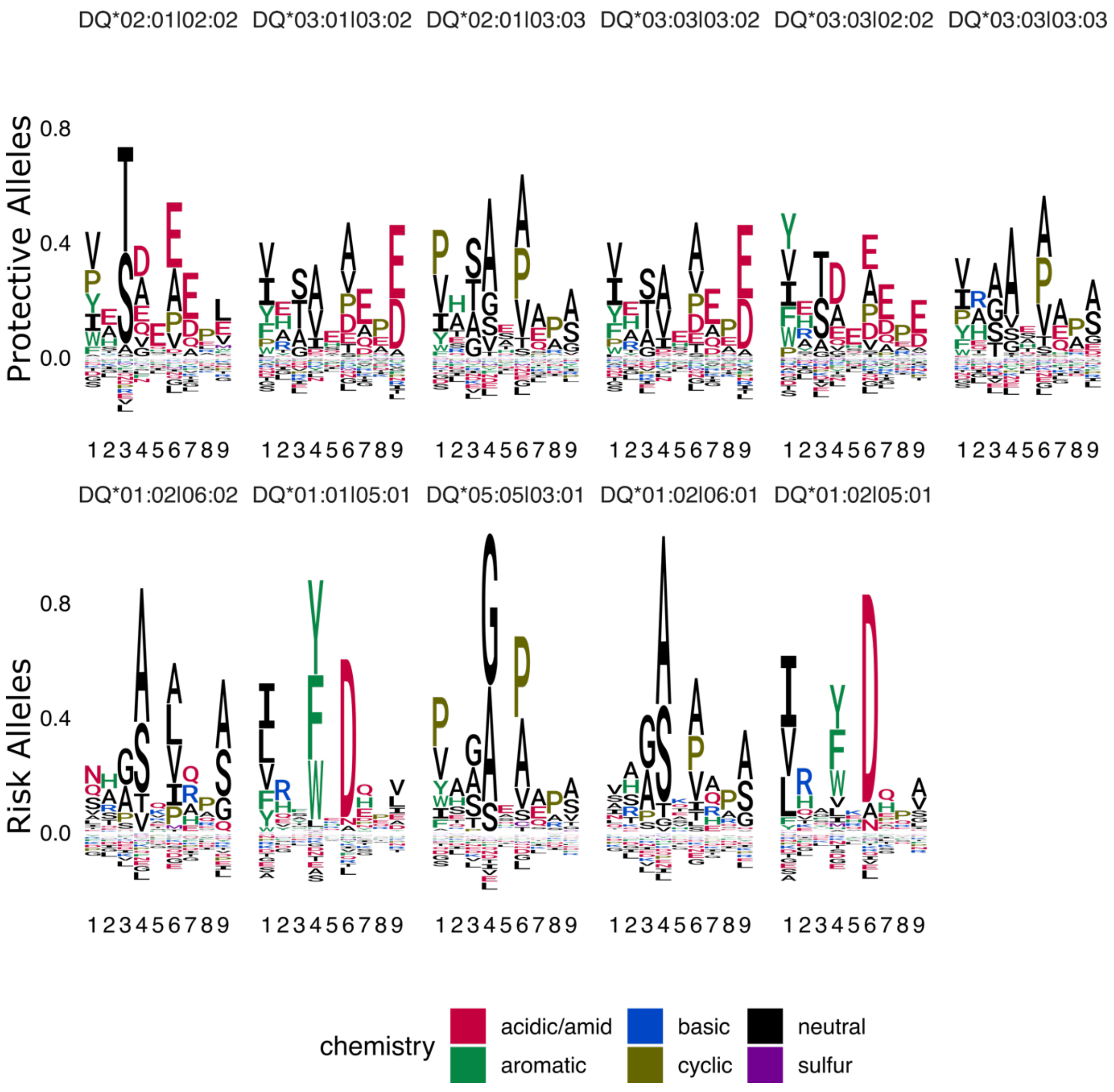

***Supplementary Figure 34:*** *Binding logo plot of associated HLA-DQ alleles. The allele names are shortened, e.g., DQA1*02:01-DQB1*02:02 is written as DQ*02:01|02:02. The motifs are based on the NetMHCIIpan-4.0 predictions of the binding cores of all peptides at least annotated as weak binders. The single letters represent the one letter amino acid code colored by the chemical properties of the amino acids.*

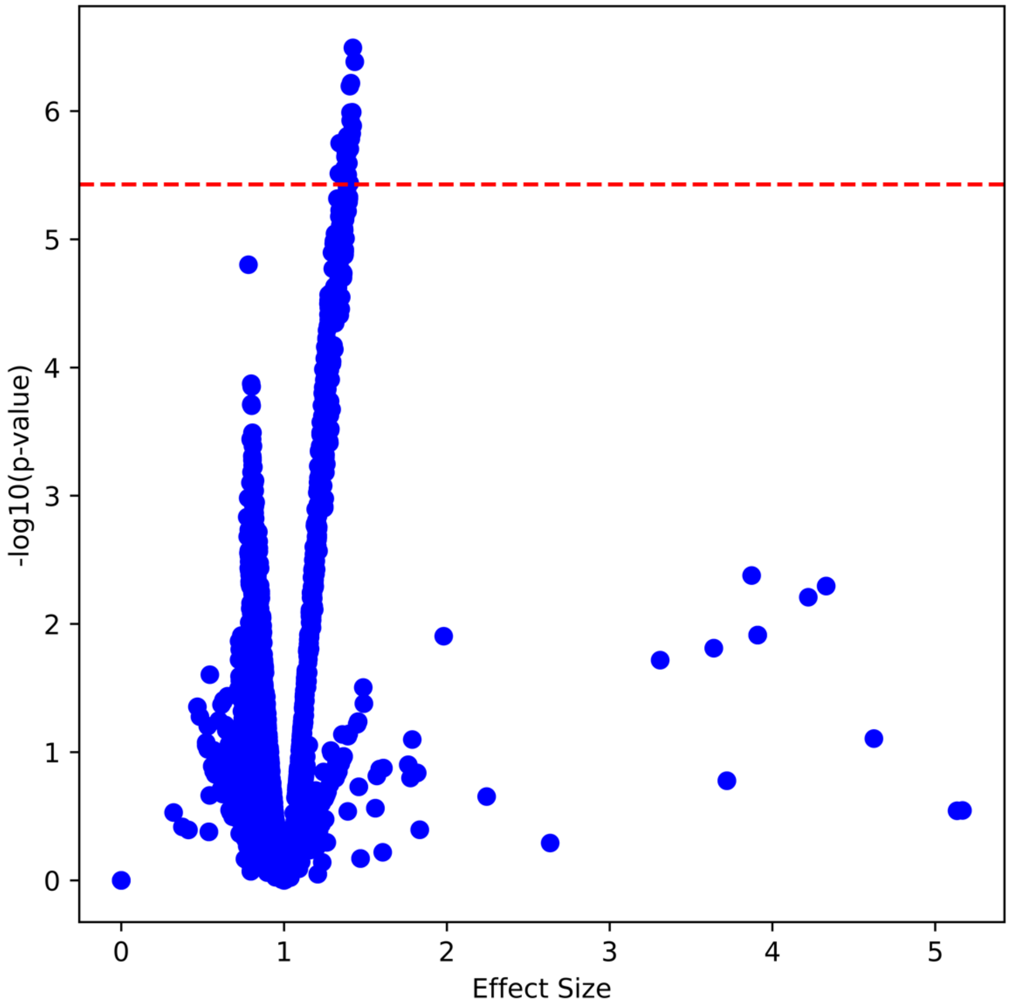

***Supplementary Figure 35:*** *Vulcano plot of the PepWAS analysis. Each dot represents the PepWAS results for one peptide. The red dotted line represents the Bonferroni corrected P-value* (P-value <3.73 × $10^{-6}$ based on 13,411 peptides).

#### Supplementary Tables

***Supplementary Table 1:*** *Sample number before, during, and after QC. Numbers in white lines represent sample numbers to be removed.*

| | UC | Control | All traits |
|---|---|---|---|
| Metadata | 950 | 4681 | 20563 |
| GSA+Exome (GSA/Exome) | | | 19769 (20554/19772) |
| GSA input | 950 | 4680 | 20554 |
| -non German samples (Metadata) | 0 | 0 | 1312 |
| -only Exome data | 0 | 0 | 3 |
| -only GSA data | 31 | 171 | 785 |
| -duplicate samples | 1 | 46 | 256 |
| -unique blacklist | 32 | 217 | 2065 |
| GSA QC input | | | **18492** |
| -Missingness outlier | 12 | 77 | 392 |
| -Heterozygosity outlier | 2 | 5 | 41 |
| -PCA outliers | | | 506 |
| -duplicates | 0 | 0 | 2 |
| -unique QC removed | 53 | 121 | 903 |
| GSA after QC | 865 | 4342 | 17589 |
| -Relatives | 2 | 157 | 1672 |
| -relatives not already removed | | | 1634 |
| GSA final Qced | 863 | 4185 | **15955** |
| Exome input | 119 | 4509 | 19772 |
| -het/hom | 3 | 11 | 43 |
| -TiTv | 0 | 0 | 1 |
| -singletons | 6 | 11 | 65 |
| -missingness | 0 | 0 | 7 |
| -sex | 0 | 0 | 13 |
| -unique QC removed | 6 | 16 | 84 |
| Exome final Qced | 913 | 4493 | **19688** |
| GSA and Exome Qced (including relatives) | | | 17138 |
| Association (no relatives, only UC and Controls) | 863 | 4185 | 5048 |

***Supplementary Table 2:*** *Genes and transcripts used to generate the proteome. Given are next to the genetic hg38 location and the ensemble-ids also the biotype of the transcript, the hgnc symbol and the uniprot gene ids.*

[See separate xlsx-file]

***Supplementary Table 3:*** *At least nominal significantly associated lead variants identified in the "imputed genotyping" dataset or the "Exome" dataset. For each locus an identifier is given in the "NR" column and the band information, further in case of a locus top hit it is annotated whether the locus is LD supported. For comparison additional entries from external sources were added. Those sources are the GWAS catalog (dataset named by the first author of the publication; namely: Liu JZ[3], de Lange K[4], Silverberg MS[5], McGovern DP[6], Jostins L[7], Barrett JC[8], Anderson CA[9], Asano K[10], Okamoto D[11], Ellinghaus D[12]) as well as information for the exact same variants or variants in high-LD ($R^2$>0.9) from the publicly available RICOPILI summary statistics (IBD_UC_1KG_oct13). The specific variant is characterized by its rs-id, the chromosomal position in*

*GRCh38 (hg38) and GRCh37 (hg19) as well as the given alleles. From the association analysis the P-value (p.value) the OR with its 95% confidence interval (CI_L95 and CI_U95) as well as the beta and standard error (SE) are given. Further, the MAPPED_TRAIT is shown, which is always ulcerative colitis for our own data and the dataset extracted from RICOPILI but varies for data from the GWAS catalog as also variants including IBD are listed in case no association with UC could be identified for this locus in the database but another IBD related trait. Further the allele frequencies separated by patients (AF.Cases) and controls (AF.Controls) is given. For the RICOPILI data the frequencies are as given by Hapmap. For imputed variants, the imputation info is given. The R2 is related to imputed genotyping lead variant, the corresponding dataset is also given in the dataset_ld column. The mapped gene and the related impact on the protein structure (change) are listed as well as gtex associated genes associated with the variant.*

[See separate xlsx-file]

***Supplementary Table 4:*** *The association results with the HLA imputed data. In the "type" column the type of the analyzed variant is given as one of the following: 1-field HLA, 2-field HLA, nuc, or prot. The HLA gene name (locus) and the exact description of the variant (name) together with the chromosome (CHR) and position in the human reference genome GRCh38 (hg38) and GRCh37 (hg19) and the position within the protein sequence in case of nucleotides (nuc) and amino acids (prot) specify the exact variant. The alleles A1 and A2 are either the one letter nucleotide or amino acid code or in the case of polymorphic variants and HLA alleles noted as present-absent (P and A). From the association analysis the P-value (p.value), the OR with its 95% confidence interval (CI_L95 and CI_U95), as well as the standard error (SE) are given. Further the minor allele frequency (MAF) as well as the allele frequency separated by cases (AF.Cases) and controls (AF.Controls) is included. Additionally, for the HLA alleles the posterior probability is listed as a reliability score of the imputation.*

[See separate xlsx-file]

***Supplementary Table 5:*** *The significant associated peptides from the PepWAS analysis (**sheet: peptides**) and the information condensed on the transcript (sheet: transcripts) and gene level (sheet: genes). For each peptide the association statistics are given as effect size (effect_size) and p-value (p) further the corresponding transcripts (ENSTs) with the position in GRCh38, the ensemble gene ids (ENSG) and the hgnc symbol is given. It is noted whether the peptide is present in the reference proteome (mutations) and in how many samples (n). The sample numbers with a specific peptide are also listed separated by cases (n_nase) and controls (n_controls). Those values are also given as frequencies (f, f_case and f_control). Also, the OR of the frequencies (OR_freq) is included and the p-value of the fisher test on the frequencies. The mutations related to the single peptides are listed: First the mutations that are needed to generate the sequence (nucchanges and protchanges), further the different amino acid positions in different transcripts are listed and then all mutations, that would change the sequence of the peptide, are listed. The column HLA lists the HLA alleles predicted to bind the peptide. The number (n_immunopeptidome_blood) and sequences (immunopeptidome_blood) of identified peptides in the 25 immunopeptidomes published in ElAbd et al.[13] are presented. On the transcript level (**sheet: transcripts**) for each ensemble transcript (ENST) the uniprot gene id and the ensemble protein id together with the length of the amino acid sequence (lengthAA) are noted. The number of peptides per transcript are given (n_hits). As the peptides are generated by a sliding window approach and peptides binding HLA class II are longer than the binding pocket, often neighboring peptides are predicted as similar good peptides and a single missense mutation might have only a small impact on the binding affinity, therefore also the number of peptides where less than 9 amino acids are in the same order are given (n_no9AAoverlap). Further, the number of PepWAS hits is separated by those with a mutation (n_hits_mut) and those present in the reference (n_hits_ref). Additional numbers of related mutations (n_relevant_mut) are given as mutations necessary to form a PepWAS hit (necessary_mut), mutations that are changing the present peptide but both peptides are PepWAS hits (possible_mut), and mutations that modify a peptide in a way that it is not predicted as PepWAS hit anymore (forbidden_mut). On the gene level (**sheet:genes**) the different transcripts are summarized, with the majority of the previously described attributes, and whether all peptides annotated to one gene are expressed within one transcript (AllHitsInOneENST). Further information about the expression of the genes is given as reported in Taman et al.[14] and in Linggi et al[15].. Further if the confidence set of associated variants includes any GTEx[16] variants the effect is given in comparison to the risk variants (risk_variant_lead_to_expression) as "decreased" or "increased" expression. The immunopeptidome data are given for the whole genes independent of the location of the PepWAS hits.*

[See separate xlsx-file]
